# Cross-validation of wearable light loggers across electric and natural light conditions: focus on illuminance, melanopic EDI, and CCT

**DOI:** 10.64898/2026.10.06.26364836

**Authors:** Peiman Pilehchi-Ha, Oliver Stefani, Marcel Schweiker, Yvonne de Kort, Karin Smolders, Jan-Frieder Harmsen

**Author notes:** **Address of correspondence:** Jan-Frieder Harmsen, Institute for Occupational, Social and Environmental Medicine, RWTH Aachen University, Pauwelsstraße 30, 52074 Aachen, Germany.

## Abstract

Wearable light loggers are increasingly used to assess light exposure in everyday environments in studies of circadian physiology and sleep, yet their accuracy under diverse lighting conditions remains insufficiently understood. This study evaluated three commonly used wearable light loggers (*ActLumus*, *Lido* and *OcuWEAR*) by comparing measurements of photopic illuminance, melanopic equivalent daylight illuminance (m-EDI), and correlated colour temperature (CCT) against a reference spectroradiometer under controlled laboratory conditions and in the field. Measurements were conducted across fluorescent, incandescent, and multiple LED sources, as well as under dynamic daylight exposure. Device performance depended strongly on both light source characteristics and metric type. Illuminance exhibited lower variability, whereas m-EDI and CCT showed substantial device- and condition-specific biases, particularly under spectrally atypical illumination conditions. These differences were consistent with spectral mismatch effects arising from standard photometric calibration practices. Importantly, the impact of measurement bias varied by exposure context: under dim light conditions, small absolute deviations led to substantial misclassification relative to currently recommended thresholds, whereas brighter electric light and daylight conditions were more robust. These findings highlight that device- and context-dependent biases can influence the interpretation of light exposure in circadian and sleep research, underscoring the need for careful device selection and data interpretation.

## Introduction

Light plays a fundamental role in human physiology through both image-forming (visual) and non-image-forming pathways. The latter are primarily mediated by intrinsically photosensitive retinal ganglion cells (ipRGCs), contribute to the regulation of circadian rhythms, sleep timing, mood, alertness, and cognitive performance ^1–11^. The discovery of ipRGCs and their melanopsin-mediated signalling provided a biological basis for the non-visual effects of light ^12–14^. These insights have motivated a shift from reporting photopic illuminance alone toward using metrics that better represent the spectral sensitivity of ipRGCs, most prominently melanopic equivalent daylight illuminance (melanopic EDI in lx). Consequently, reporting melanopic EDI alongside photopic illuminance (lx) and correlated colour temperature (CCT, K) is now increasingly standard in studies addressing circadian, sleep, and health outcomes ^15,16^.

Historically, studies examining the effects of light on sleep and circadian physiology were largely confined to laboratory environments using high-precision spectroradiometers ^17^. Such experimental work has established the causal influence of morning light exposure on circadian phase advancement, the phase-delaying effects of evening light, and the acute alerting effects of light across the day ^18^. Yet laboratory conditions do not capture the variability of everyday light environments, which are shaped by behavioural patterns, indoor lighting practices, and cultural or occupational factors ^15,19–22^. These limitations have increased the need for field-based studies that capture personal light exposure under everyday living conditions. Wearable light loggers make such measurements possible by continuously recording individuals’ light exposure in their natural environments. Recent field studies have demonstrated the value of this approach. For example, Didikoglu et al. (2023) used an open-source wrist-worn light logger to monitor melanopic EDI exposure, sleep timing, and subjective sleepiness across seven days in adults living their normal daily lives ^23^. They found widespread deviation from expert consensus-based recommendations for melanopic light exposure levels, which are informed by experimental evidence on human circadian, neuroendocrine, and alerting responses to ocular light and recommend ≥250 lx melanopic EDI during daytime, ≤10 lx during the 3 h before bedtime, and ≤1 lx during sleep ^24^. Specifically, real-world measurements indicate insufficient daytime light exposure alongside substantial evening and nighttime melanopic light exposure ^25^. Consistent with these recommendations, lower morning melanopic EDI was associated with greater daytime sleepiness, whereas higher pre-bedtime melanopic EDI was associated with longer sleep-onset latency ^23^. These findings highlight how light exposure under everyday living conditions is associated with sleep and alertness, and demonstrate the need for accurate, well-characterised wearable devices when evaluating associations between light, sleep, and health outcomes. Reliable and valid light measurement in everyday environments is therefore important for understanding how daily light exposure patterns relate to sleep timing, circadian alignment, and subjective alertness.

Recent advances in wearable light loggers enable continuous personal light measurement in naturalistic settings, offering new opportunities to quantify individuals’ “spectral diets” over time ^17,26,27^. However, the accuracy and comparability of such devices remain a critical challenge. Wearable light loggers can differ in sensor spectral sensitivity, angular response, calibration procedures, sampling intervals, and form and wearing position. Because many devices are worn on the wrist or torso rather than near the eye, they may systematically underestimate corneal light exposure ^21,28^. This is particularly important under dim lighting conditions commonly occurring during the evening and at night, where relatively small absolute measurement errors can translate into substantial relative errors and consequently affect whether exposure is classified as meeting recommended light levels. Near-corneal devices, while more accurate, may compromise comfort and compliance, influencing data completeness ^16,21^. In addition, measurement accuracy can vary across light conditions. Spectral composition and light intensity differ across daylight, indoor diffuse illumination, and LED-based light sources, which can introduce device-specific biases, especially when computing melanopic metrics ^17^.

To ensure that wearable-based field measurements translate into meaningful physiological interpretations, it is essential to systematically validate wearable light loggers against spectroradiometric reference instruments under controlled lighting conditions. Therefore, the present study aims to cross-validate the performance of multiple wearable light logging devices under varying spectral composition and intensity conditions (e.g., daylight and electric light sources). We focus on illuminance, CCT, and melanopic EDI, arguably the three core metrics used in circadian and lighting research. By characterising device accuracy across representative lighting conditions, this study provides methodological guidance for interpreting wearable-based field data and designing future research that links light exposure in everyday environments to sleep, circadian health, and well-being.

## Results

### Correlation between two ActLumus devices

To assess the consistency between two units of the same light wearable logger model, we compared measurements obtained from two *ActLumus* devices placed side-by-side (Figure 1 and 2B) under identical lighting conditions during the COLORBOX and FLUORESCENT light conditions. Given that both devices were calibrated by the respective manufacturers and were expected to report comparable values under equal exposure, this analysis aimed to evaluate the extent to which sensor position (e.g., as the three wearable light loggers of interest could not be placed in the same three-dimensional position relative to the light source) might affect measured light levels. Figure S1 presents the correlations between the two devices for photopic illuminance (lx) and melanopic EDI (lx) across the full dataset. Both measures demonstrated a very strong linear relationship, with coefficients of determination of R² = 0.996 for photopic illuminance and R² = 0.999 for melanopic EDI (Figure S1). The corresponding mean absolute differences between the two devices were 24 lx for illuminance and 11 lx for melanopic EDI. The data points closely align with the identity line, indicating only minor deviations between the two sensors. Therefore, for the following experimental conditions, we applied only one *ActLumus* sensor and assumed that having the three different wearable light loggers placed directly next to each other on a horizontal line (Figure 1 and 2B) relative to the light source does not induce meaningful differences for cross-validation purposes.

**Figure 1:**
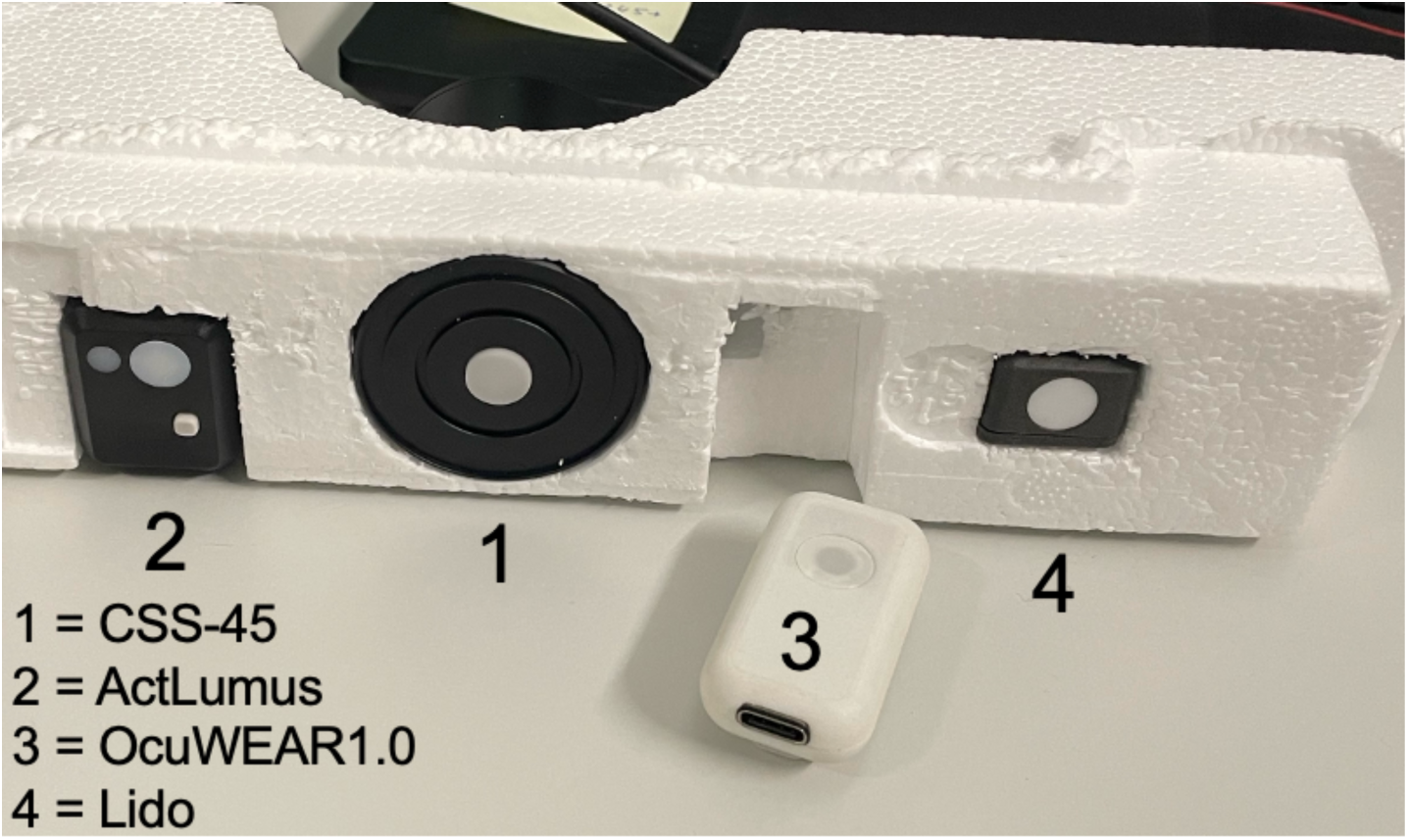
The three light loggers, as well as the reference spectroradiometer, were placed next to each other horizontally in a custom-designed styrofoam testbed. This basic arrangement was used for all cross-validation experiments.

**Figure 2:**
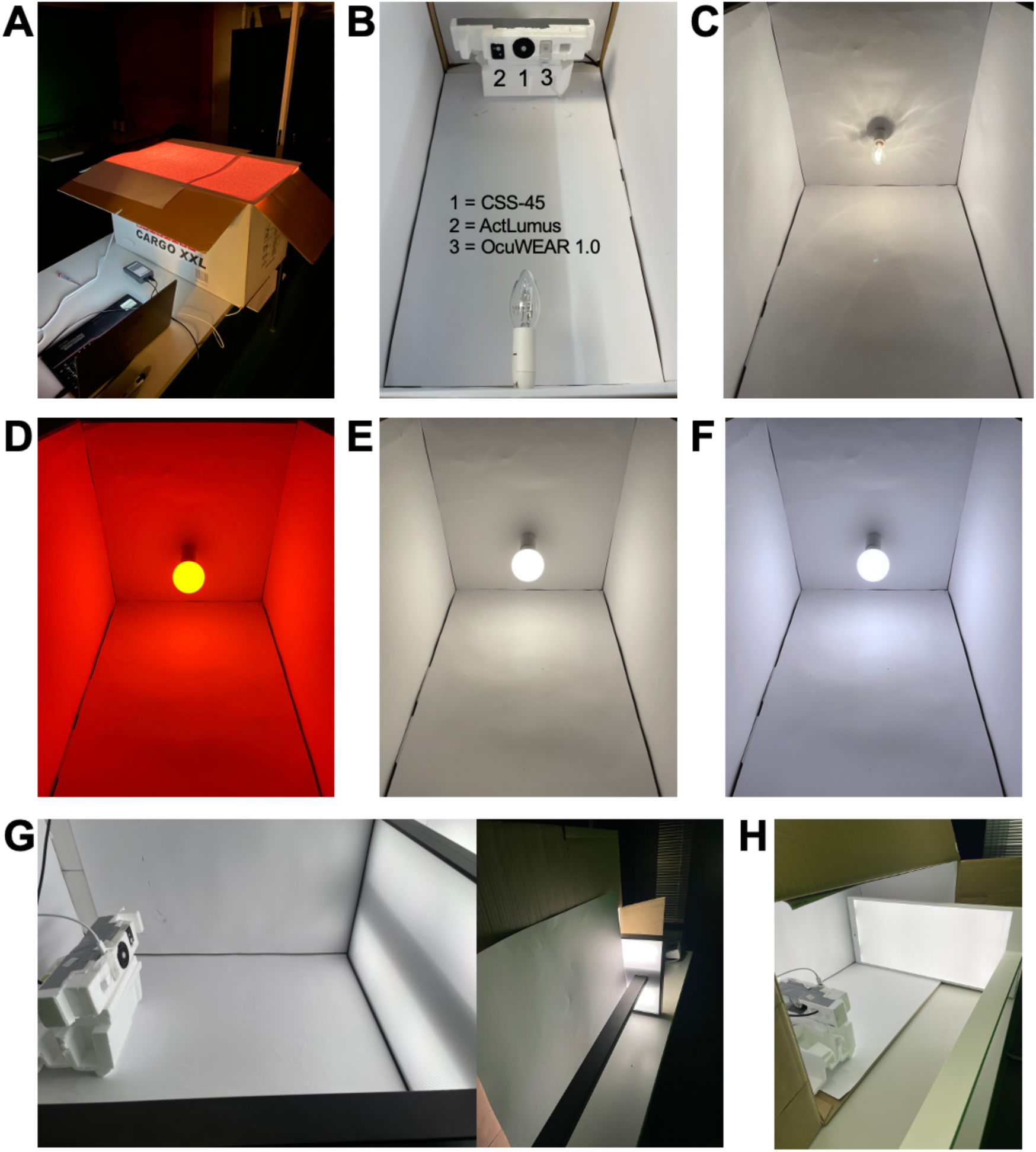
Experimental set-up to test INCANDESCENT and different LED light sources. A custom-made cardboard box covered with white glossy photo paper served as a closed light box (A), in which different electric light sources were installed to face the wearable light loggers (ActLumus and OcuWEAR) and the reference spectroradiometer at approximately 30-40 cm distance, depending on the light source (B). An incandescent light bulb (C) and the Vitae LED light bulb in three different modes (Orange (D), Warm White (E) and White (F)) were installed through a small hole in the cardboard box. Two standing lamps were further tested, put down lying on their side (Daylight-mimicking Spectrasol Terra LED lamp (G) and a TRILUX LuceoS Act S LED standing lamp (H)) with the downward-facing light directed to the measurement set-up, while the cardboard box was also turned on its side.

To interpret the following results, a positive bias indicates that the wearable device overestimates values relative to the reference spectroradiometer (device > CSS), whereas a negative bias indicates underestimation (device < CSS).

### Performance in the FLUORESCENT light scenarios

Under *FLUORESCENT* lighting conditions, all three wearable light loggers exhibited systematic deviations from the reference spectroradiometer, with clear device-specific patterns (Table 1; Figure 3).

**Figure 3:**
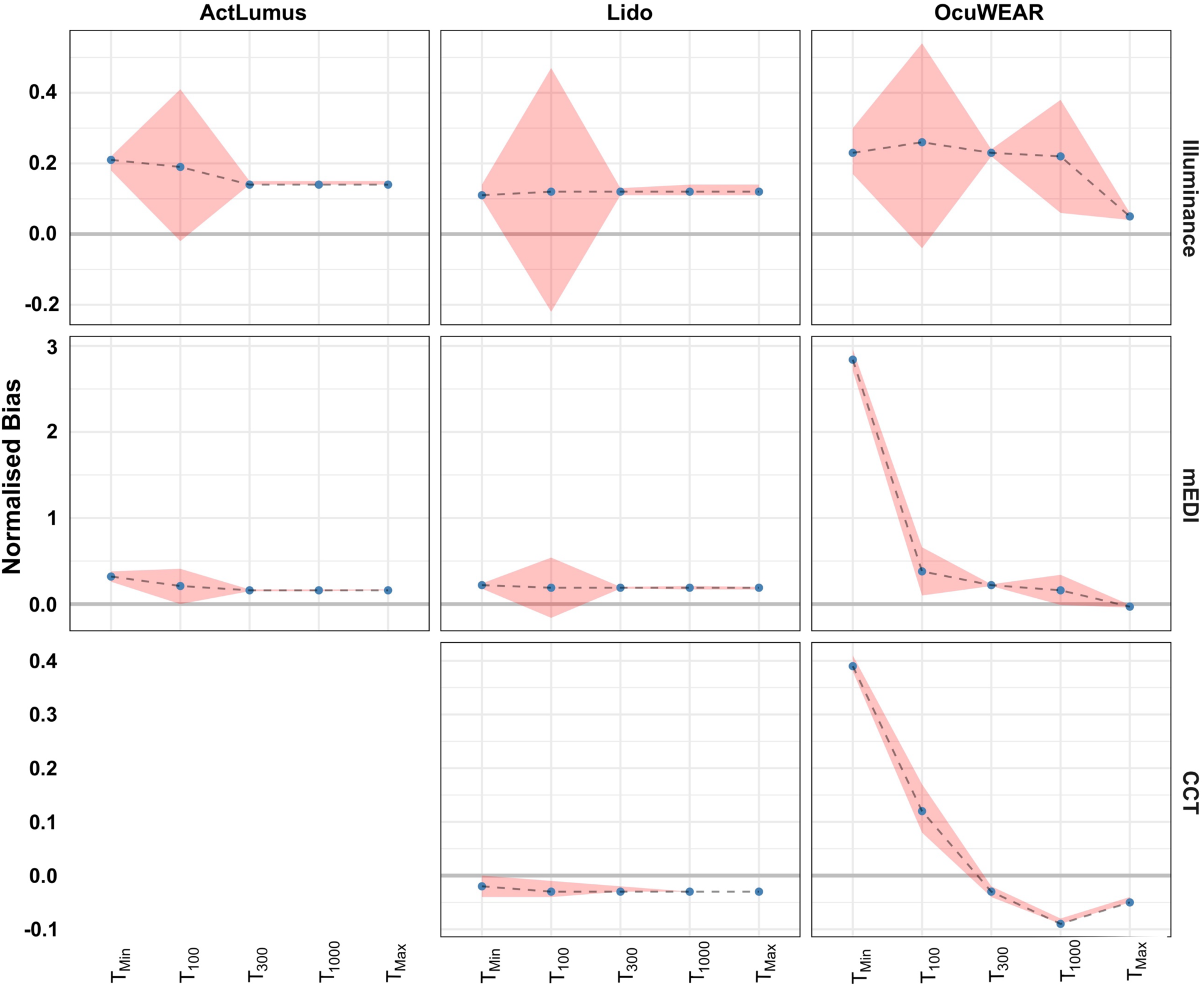
Bland-Altman normalised bias and LoA for light loggers across light targets under FLUORESCENT light. T_max_ was 2004 lx measured using CSS-45.

**Table 1:** Bland-Altman result for light loggers exposed to the FLUORESCENT light source. Tmax was measured by CSS-45, equal to 2004 lx. A positive bias value indicates that the respective light logger device displayed higher values compared to the reference spectroradiometer (overestimation), and vice versa for negative values (underestimation).

| Light logger | Metric | Target condition | Bias | Lower LoA | Upper LoA |
| --- | --- | --- | --- | --- | --- |
| *ActLumus* | Illuminance (lx) | T<sub>10</sub> | 3 | 2 | 3 |
|  |  | T<sub>100</sub> | 20 | 19 | 21 |
|  |  | T<sub>300</sub> | 47 | 44 | 50 |
|  |  | T<sub>1000</sub> | 159 | 150 | 168 |
|  |  | T<sub>Max</sub> | 284 | 270 | 298 |
|  | Melanopic EDI (lx) | T<sub>10</sub> | 2 | 1 | 2 |
|  |  | T<sub>100</sub> | 13 | 12 | 14 |
|  |  | T<sub>300</sub> | 31 | 29 | 33 |
|  |  | T<sub>1000</sub> | 105 | 100 | 111 |
|  |  | T<sub>Max</sub> | 189 | 180 | 198 |
| *Lido* | Illuminance (lx) | T<sub>10</sub> | 2 | 1 | 2 |
|  |  | T<sub>100</sub> | 14 | 13 | 15 |
|  |  | T<sub>300</sub> | 41 | 38 | 44 |
|  |  | T<sub>1000</sub> | 138 | 123 | 154 |
|  |  | T<sub>Max</sub> | 251 | 231 | 270 |
|  | Melanopic EDI (lx) | T<sub>10</sub> | 1 | 1 | 1 |
|  |  | T<sub>100</sub> | 13 | 12 | 14 |
|  |  | T<sub>300</sub> | 37 | 36 | 39 |
|  |  | T<sub>1000</sub> | 127 | 117 | 136 |
|  |  | T<sub>Max</sub> | 221 | 209 | 233 |
|  | CCT (K) | T<sub>10</sub> | -60 | -118 | -1 |
|  |  | T<sub>100</sub> | -106 | -106 | -106 |
|  |  | T<sub>300</sub> | -104 | -104 | -104 |
|  |  | T<sub>1000</sub> | -107 | -107 | -107 |
|  |  | T<sub>Max</sub> | -110 | -110 | -110 |
| *OcuWEAR* | Illuminance (lx) | T<sub>10</sub> | 3 | 2 | 4 |
|  |  | T<sub>100</sub> | 26 | 24 | 28 |
|  |  | T<sub>300</sub> | 76 | 73 | 80 |
|  |  | T<sub>1000</sub> | 249 | 75 | 422 |
|  |  | T<sub>Max</sub> | 102 | 86 | 119 |
|  | Melanopic EDI (lx) | T<sub>10</sub> | 15 | 14 | 15 |
|  |  | T<sub>100</sub> | 25 | 23 | 26 |
|  |  | T<sub>300</sub> | 44 | 41 | 46 |
|  |  | T<sub>1000</sub> | 109 | -4 | 221 |
|  |  | T<sub>Max</sub> | -32 | -44 | -21 |
|  | CCT (K) | T<sub>10</sub> | 1164 | 1121 | 1207 |
|  |  | T<sub>100</sub> | 521 | 469 | 573 |
|  |  | T<sub>300</sub> | -114 | -138 | -90 |
|  |  | T<sub>1000</sub> | -346 | -368 | -324 |
|  |  | T<sub>Max</sub> | -190 | -205 | -175 |

*ActLumus* showed a consistent positive bias for both illuminance and melanopic EDI across all target conditions, indicating systematic overestimation relative to the reference. The magnitude of this overestimation increased with light intensity, and the limits of agreement widened proportionally as light levels increased. *Lido* demonstrated a similar positive bias in illuminance and melanopic EDI to *ActLumus* across all target conditions. Bias magnitude increased with target level for both metrics. In contrast, CCT measurements from *Lido* consistently showed a small negative bias across all target conditions, indicating a slight but systematic underestimation of colour temperature under fluorescent spectra.

*OcuWEAR* exhibited more complex and target-dependent behaviour. While illuminance and melanopic EDI generally showed positive bias at lower and intermediate light levels (i.e., overestimation), both the magnitude and direction of bias varied at higher intensities. In particular, melanopic EDI shifted to a negative bias at maximum light levels, indicating underestimation under these conditions. Illuminance measurements showed substantially wider limits of agreement at intermediate targets, suggesting reduced precision. CCT measurements also transitioned from positive bias at low light levels (overestimation) to negative bias at higher intensities (underestimation), indicating non-monotonic and spectrum-dependent performance.

These patterns are reflected in the normalised Bland–Altman analysis (Figure 3), where *ActLumus* demonstrates relatively stable and near-zero bias across targets, Lido shows moderate target-dependent variability, and *OcuWEAR* exhibits pronounced variability and wider limits of agreement across all metrics.

### INCANDESCENT performance

Under INCANDESCENT lighting conditions, both *ActLumus* and *OcuWEAR* exhibited target-dependent deviations from the reference spectroradiometer, with differences in the direction and magnitude of bias across light levels (Table S1; Figure 4).

**Figure 4:**
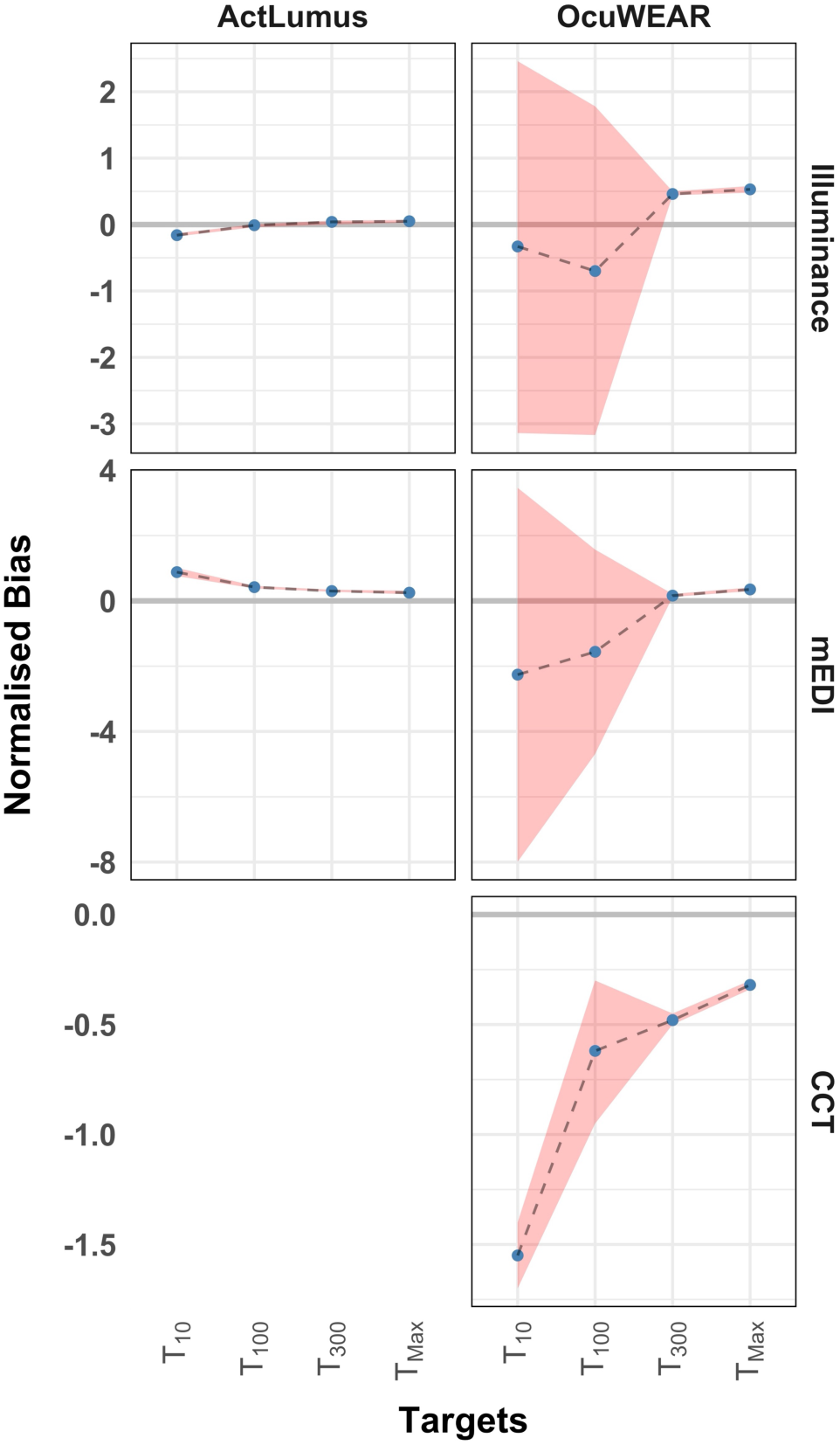
Bland-Altman normalised bias and LoA for light loggers across light targets under an incandescent lamp. T_max_ was 626 lx measured by CSS.

For *ActLumus*, illuminance showed a negative bias at lower target conditions (underestimation) and a positive bias at higher targets (overestimation). Melanopic EDI exhibited a consistent positive bias across all target conditions, indicating systematic overestimation relative to the reference. The magnitude of bias increased with light intensity for both metrics, and the limits of agreement also increased with target level.

For *OcuWEAR*, illuminance showed a negative bias at lower target conditions (underestimation) and a positive bias at higher targets (overestimation). Melanopic EDI followed a similar pattern, with negative bias at lower targets and positive bias at higher light levels. CCT measurements showed a consistent negative bias across all target conditions (underestimation of CCT). Limits of agreement for both illuminance and melanopic EDI were wider at lower light levels and became narrower with increasing target level.

### LED performance

#### COLORBOX

Under COLORBOX lighting conditions, all three wearable light loggers showed device- and spectrum-dependent deviations from the reference spectroradiometer, with distinct patterns between the green (T_G_ with light intensity of photopic 100 lx) and red (T_R_ with similar light intensity) light settings. The reference spectroradiometer (*CSS-45*) did not provide CCT outputs for these selected light conditions; therefore, Bland–Altman analyses were restricted to illuminance and melanopic EDI metrics. Absolute values are presented in Table S2.

*ActLumus* exhibited a consistent positive bias and overestimation in illuminance under both T_G_ and T_R_ conditions. For melanopic EDI, bias direction differed between conditions, with a small positive bias under T_G_ (small overestimation) and a negative bias under T_R_ (underestimation). *Lido* showed condition-dependent behaviour. Illuminance measurements exhibited a positive bias under T_G_ (overestimation) and values close to zero bias under T_R_. In contrast, melanopic EDI consistently showed positive bias under both conditions (overestimation) with larger deviations observed under T_R_ compared to T_G_.

*OcuWEAR* demonstrated pronounced spectrum-dependent variability. Illuminance measurements showed a positive bias under T_G_ and a negative bias under T_R_. For melanopic EDI, the pattern was reversed.

These patterns are reflected in the normalised Bland–Altman analysis (Figure 5), where *ActLumus* shows relatively small variation between conditions, *Lido* shows condition-dependent shifts, and *OcuWEAR* exhibits stronger variation between T_G_ and T_R_ across both metrics.

**Figure 5:**
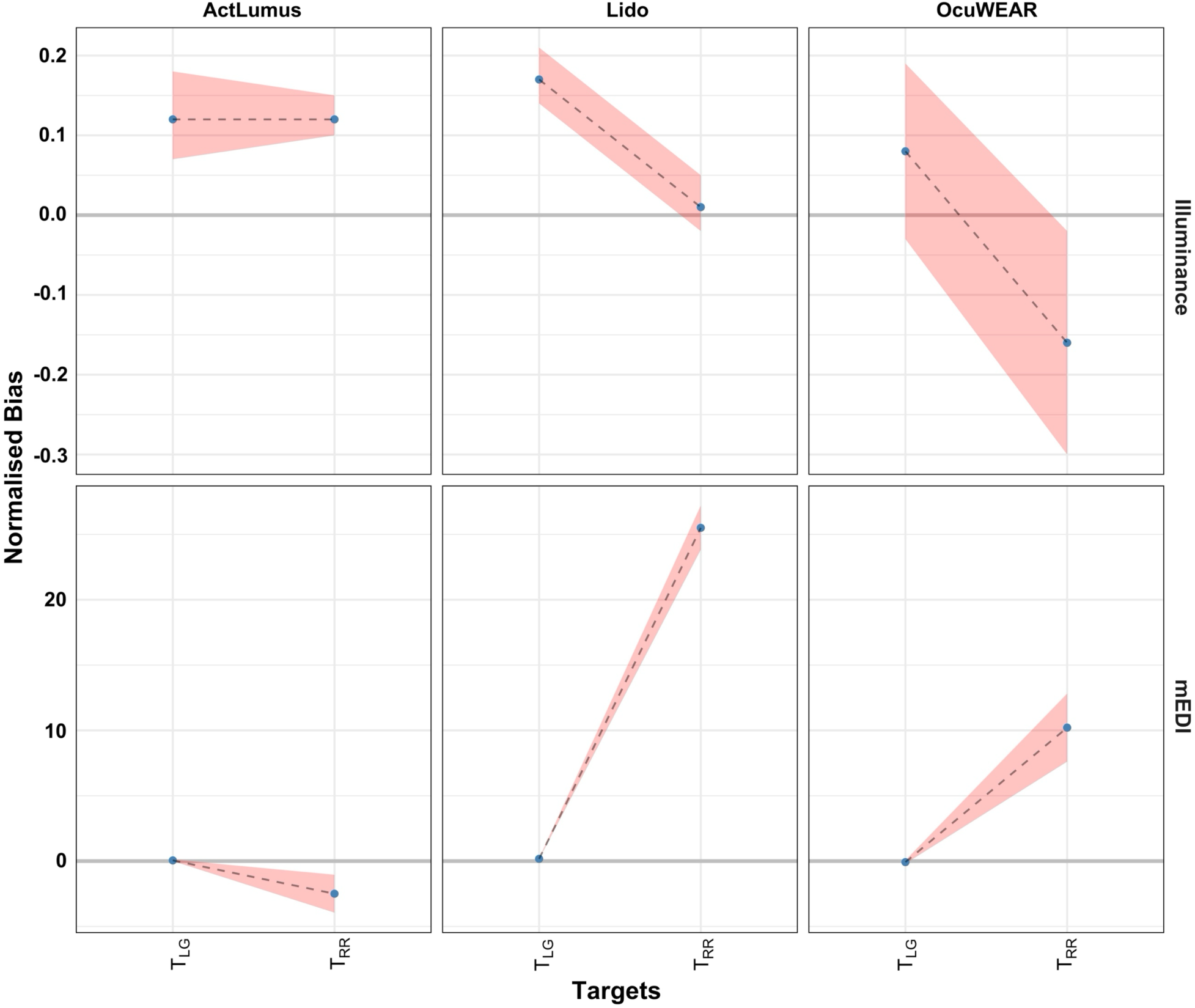
Bland-Altman normalised bias and LoA for light loggers across two light conditions in the COLORBOX. T_G_ is Light Green (102 lx) and T_R_ is Red (104 lx).

##### Standing daylight-mimicking lamp (SDL)

Under SDL, both *ActLumus* and *OcuWEAR* showed systematic deviations from the reference spectroradiometer, with device- and target-dependent patterns (Table S3; Figure 6). *ActLumus* exhibited a consistent positive bias for both illuminance and melanopic EDI across all target conditions. The magnitude of this overestimation remained relatively stable across targets, with only minor variation, and the limits of agreement decreased with light intensity.

**Figure 6:**
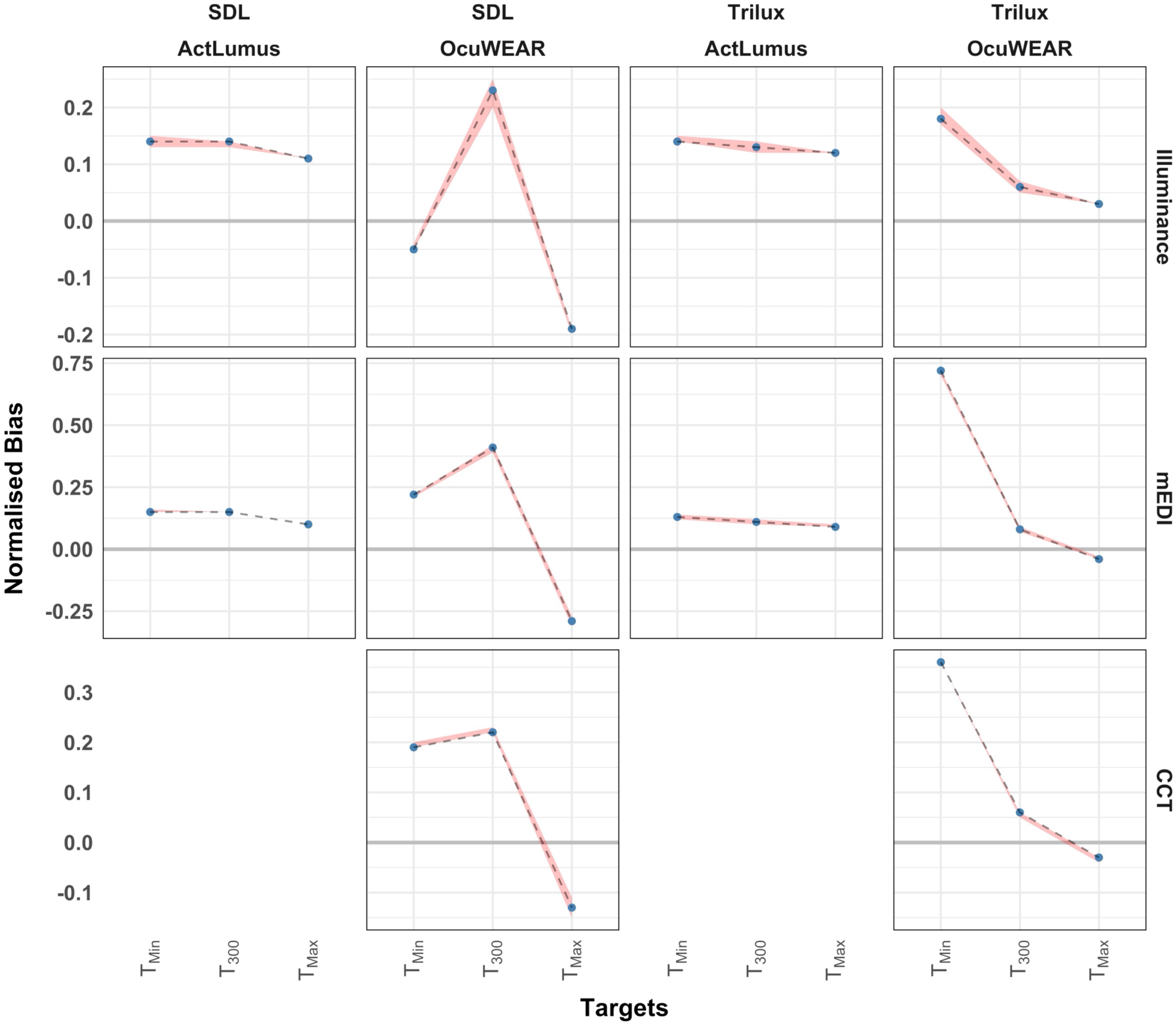
Bland-Altman normalised bias and LoA for light loggers across light targets under light sources of Spectrasol Daylight (SDL) and Trilux light lamp. For Trilux, T_Min_ and T_Max_ were 46 lx and 4299 lx, respectively, measured by CSS. For SDL, T_Min_ and T_Max_ were 133 and 12759 lx, respectively, measured by CSS.

*OcuWEAR* showed non-monotonic, target-dependent behaviour. For both illuminance and melanopic EDI, bias increased from negative or near-zero values at the lowest target to a clear positive peak at the intermediate target, followed by a strong shift to negative bias at the highest target, indicating underestimation at high light levels. This pattern reflects a reversal in bias direction across the tested range. Limits of agreement widened at intermediate intensities. CCT measurements for *OcuWEAR* followed a similar target-dependent pattern, with positive bias at lower targets and a transition toward near-zero and negative bias at higher intensities. These patterns are reflected in the normalised Bland–Altman analysis (Figure 6), where *ActLumus* shows relatively small variation across targets. At the same time, *OcuWEAR* exhibits stronger target-dependent variation and wider limits of agreement across all metrics.

##### LED standing lamp (Trilux)

Under Trilux, both wearable light loggers showed systematic deviations from the reference spectroradiometer, with distinct target-dependent patterns (Table S4; Figure 6). *ActLumus* exhibited a consistent positive bias for both illuminance and melanopic EDI across all target conditions. The magnitude of this overestimation showed only minor variation across targets, and the limits of agreement increased with light intensity.

*OcuWEAR* also showed positive bias for illuminance across all target conditions, with decreasing bias magnitude from low to high light levels. In contrast, melanopic EDI exhibited a target-dependent change in bias direction, with positive bias at lower and intermediate targets and negative bias at the highest target. CCT measurements for *OcuWEAR* showed a strong positive bias at the lowest target, followed by a reduction toward near-zero bias at intermediate levels and a slight negative bias at the highest target, indicating a progressive shift in bias across target conditions.

These patterns are reflected in the normalised Bland–Altman analysis (Figure 6), where *ActLumus* shows relatively small variation across targets. At the same time, *OcuWEAR* exhibits target-dependent changes in the magnitude and direction of bias, particularly for melanopic EDI and CCT.

##### TUNABLE LED light bulb (Vitae)

Systematic deviations of both wearable light loggers under Vitae are presented in Table S5 and Figure 7. *ActLumus* exhibited a consistent positive bias for illuminance across all settings. The magnitude of this overestimation increased from the Orange to the White setting. Melanopic EDI showed minimal bias under the Orange setting, while positive bias (overestimation) was observed under the Warm White and White settings, with increasing magnitude at higher light levels.

**Figure 7:**
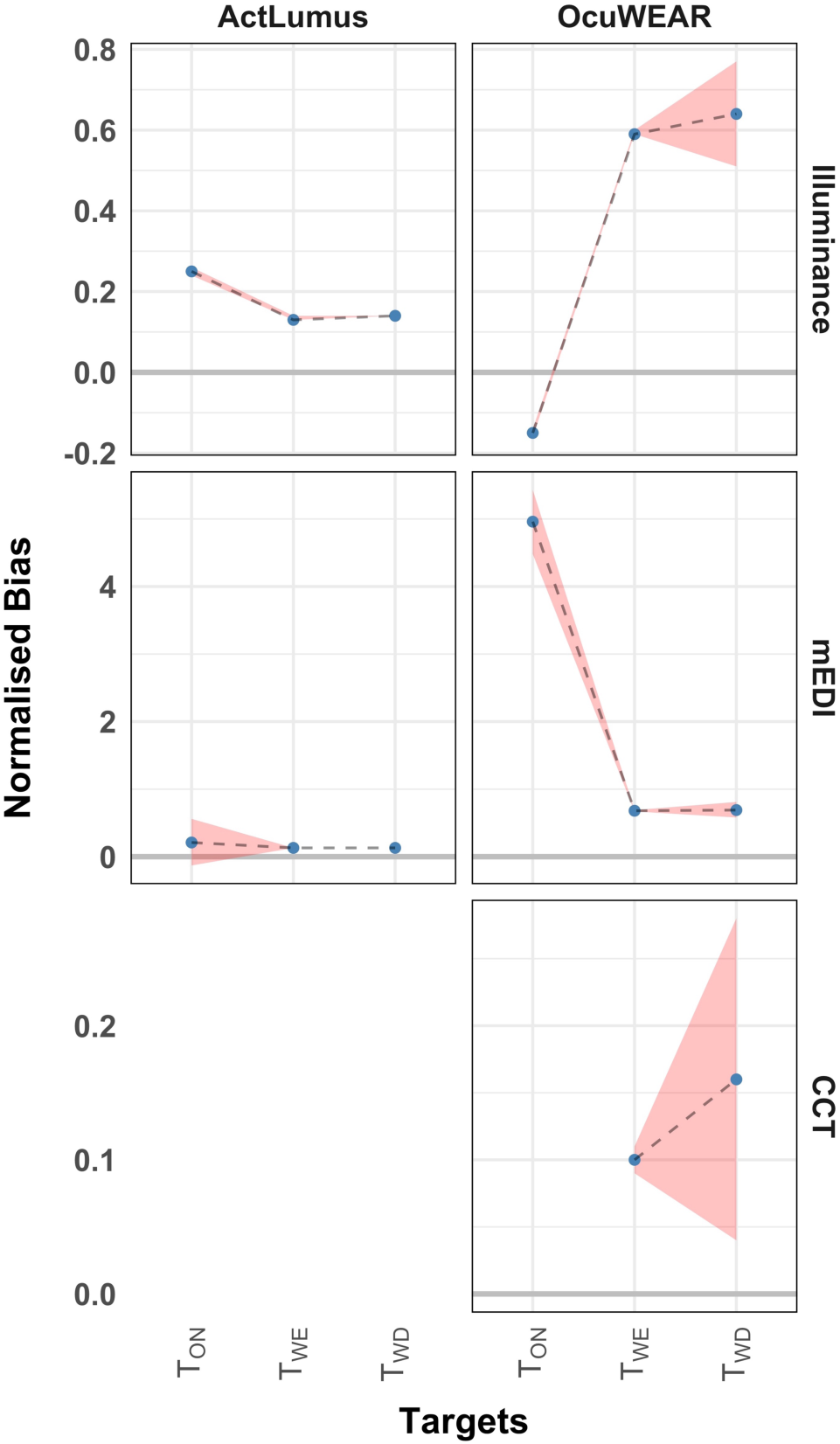
Bland-Altman normalised bias and LoA for light loggers across light conditions provided by the Vitae light bulb. T_orange_, T_warmwhite_ and T_white_ were 57 lx, 1477 lx and 2006 lx, respectively, measured by CSS.

*OcuWEAR* showed strong setting-dependent variability. Illuminance measurements exhibited a negative bias under the Orange setting, followed by a large positive bias under the Warm White and White settings, with substantially increased magnitude at higher light levels. A similar pattern was observed for melanopic EDI, with a small bias under the Orange setting and a strong positive bias under the Warm White and White settings. CCT measurements for *OcuWEAR* showed a consistent positive bias across the measured settings, with larger deviations observed under the White setting compared to Warm White.

These patterns are reflected in the normalised Bland–Altman analysis (Figure 7), where *ActLumus* shows relatively small variation across settings. At the same time, *OcuWEAR* exhibits strong setting-dependent variation, particularly for illuminance and melanopic EDI.

### Daylight performance

Figure 8 presents the temporal profiles of illuminance, melanopic EDI, and CCT measured simultaneously by the four devices of *OcuWEAR*, *Lido*, *ActLumus*, and *CSS* over the monitoring period from 09:00 to 15:00 h. Illuminance (Figure 8A) increased steadily from morning values of approximately 100 lx to peaks exceeding 10,000 lx during midday. All devices followed a similar trajectory, with minor deviations during brief fluctuation periods. Differences between devices were generally small, although *OcuWEAR* consistently reported slightly higher values than *CSS* and *Lido*, particularly during high-intensity periods. *ActLumus* showed larger divergence during a short midday peak (∼14:40), where its readings rose sharply compared with the other devices. Melanopic EDI (Figure 8B) showed a pattern closely matching the illuminance profile, as expected given the inherent relationship between photopic illuminance and melanopic EDI under daylight conditions. Values ranged from ∼100 lx m-EDI in the early morning to over 10,000 lx m-EDI at midday. As with illuminance, *OcuWEAR* tended to report slightly higher values, while the midday spike in *ActLumus* measurements was again evident. CCT (Figure 8C) started at elevated levels in the early morning (>10,000 K), dropped gradually towards a stable range of ∼6,000–8,000 K during midday, and rose again in the late afternoon. While overall trends were consistent across devices, notable differences were observed: *Lido* reported consistently higher CCT in the morning compared with *CSS* and *OcuWEAR*. Short-term fluctuations were more pronounced in *OcuWEAR* measurements.

**Figure 8:**
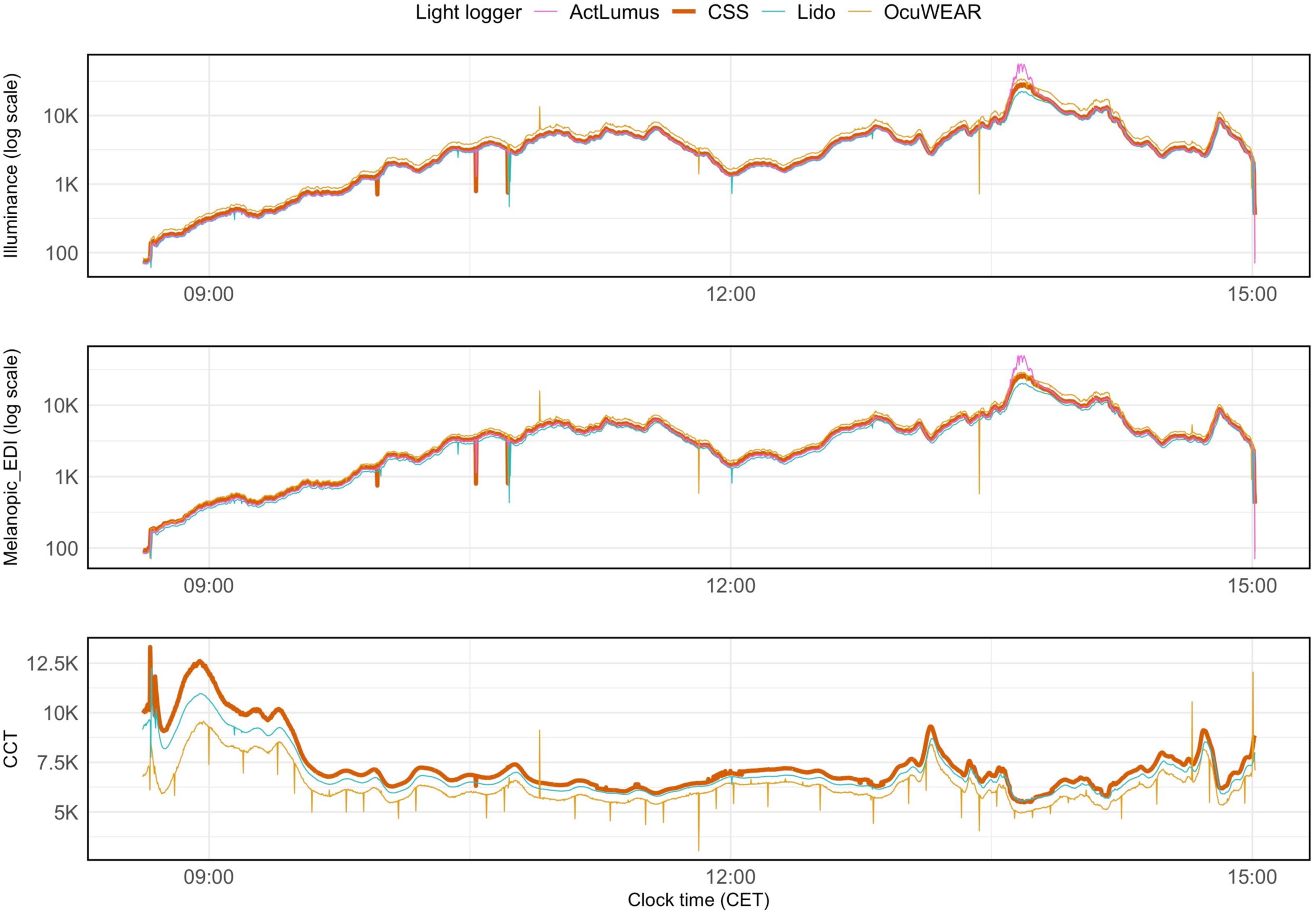
Temporal profiles of illuminance, melanopic EDI, and CCT measured by four devices during natural daylight.

Figure 9 presents the LOESS-smoothed Bland–Altman plots with moving-window LoA. Each row corresponds to one metric, while the first three columns display pairwise comparisons between *CSS* and each device (*Lido*, *OcuWEAR*, *ActLumus* for illuminance and melanopic EDI; *Lido* and *OcuWEAR* for CCT). The fourth column summarises mean bias trends for all devices. At illuminance values between 100 and 300 lx, the mean bias for all devices remained within approximately ±150 lx. Bias divergence began between 300 and 1000 lx, with *ActLumus* remaining close to zero and *Lido* and *OcuWEAR* starting to deviate in opposite directions. Above 2000 lx, the bias magnitude increased substantially. *Lido* exhibited a negative bias reaching −1000 lx for exposures more than 10000 lx. *OcuWEAR* showed a positive bias that reached approximately 2500 lx for exposure to daylight of 10000 lx, and *ActLumus* showed minimal negative bias up to approximately 10000 lx, followed by a sharp increase beyond it, reaching more than 5000 lx at higher illuminance levels. LoA remained relatively narrow (within approximately ±300–500 lx) below 1000 lx, began to widen between 1000 and 2000 lx, and became the widest above 3000 lx.

**Figure 9:**
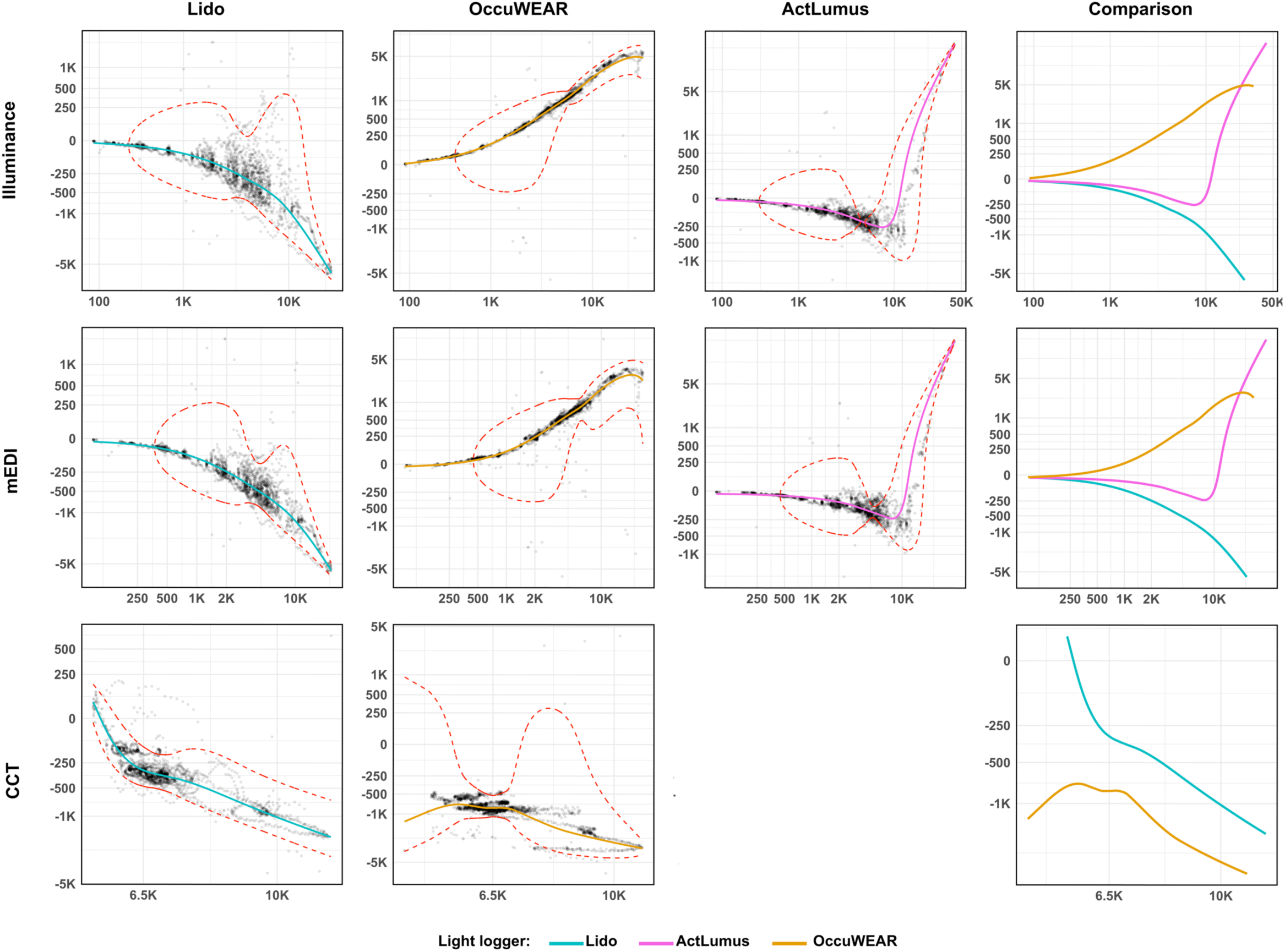
LOESS-smoothed Bland–Altman analyses with moving-window LoA for illuminance, melanopic EDI, and CCT. Axes are displayed using a pseudo-logarithmic transformation to allow simultaneous visualisation of low and high measurement ranges.

For CCT, across the measured range, mean bias was predominantly negative for both *Lido* and *OcuWEAR*, with device-specific differences occurring between clearly defined CCT intervals. Between 3000–4500 K, no data were available for *Lido*, whereas *OcuWEAR* exhibited a negative bias that decreased in magnitude with increasing CCT, ranging approximately from −1200 K to −750 K. Between 4500–6000 K, the bias trajectories differed: *OcuWEAR* shifted toward less negative bias, remaining within approximately −750 to −1000 K, while *Lido* continued to decrease, reaching approximately −300 K. Between 6500–8000 K, both devices showed increasingly negative bias. In this range, *Lido* reached approximately −300 to −500 K, while *OcuWEAR* reached approximately −800 to −1200 K. At the upper end of the range, between 8500 and 10000 K, mean bias reached approximately −500 to −1000 K for *Lido* and approximately −1200 to −3200 K for *OcuWEAR*. LoA was widest between 3500 and 4500 K for *OcuWEAR*, within approximately ±2000 K. Between 4500 and 6500 K, LoA narrowed, extending to approximately ±250-500 K. Above 7000 K, LoA were wide, with the lower limit reaching approximately −250 to −2000 K, while the upper limit remained −300 to −2250 K.

### Explorative analysis of quantifying infrared light across different light sources

In addition to illuminance and melanopic EDI, the *ActLumus* device also provides a quantification of infrared light (≥ 850 nm). Therefore, we calculated the mean and standard deviation of infrared light radiation levels in mW/cm^2^ across the different light sources and different intensities to explore if the *ActLumus* can detect expected differences, particularly between daylight, INCANDESCENT, LED and FLUORESCENT light sources. Summary statistics are provided in Table S6. Accordingly, the *ActLumus* device detected clear differences between daylight (infrared light intensity range: 0.01 to 27.8 mW/cm^2^), INCANDESCENT (0.5 to 3.3 mW/cm^2^), LED light sources (<0.6 mW/cm^2^) and FLUORESCENT light (<0.07 mW/cm^2^). As expected, there was a clear pattern of increasing infrared light intensity from the first to the second run of 5-minute data-collection trials under INCANDESCENT light conditions.

## Discussion

This study systematically evaluated the accuracy of multiple wearable light loggers under controlled electrical lighting conditions spanning a wide range of illuminance levels and spectral composition, as well as under natural dynamics of daylight.

For *ActLumus*, mean absolute biases were approximately 11.8% for illuminance and remained below 25% across all tested conditions, with a relatively narrow agreement range (mean limits of agreement ≈ 5.2%) (Table S7). Similarly, *Lido* showed stable performance for illuminance, with mean absolute biases around 11% and maximum deviations below 17%. Under *FLUORESCENT* conditions, both devices maintained comparable accuracy for illuminance (≈ 9.6–11.8%) and melanopic EDI (≈ 19–20%) (Table S8). For CCT, *Lido* exhibited consistently low deviations, with a mean absolute bias of 2.8% and maximum deviations below 3%, indicating robust chromatic performance under *FLUORESCENT* lighting. In contrast, performance diverged substantially for melanopic EDI, particularly under daylight and spectrally diverse electric light sources.

While *ActLumus* maintained moderate deviations (mean absolute bias ≈ 30%, see Table S7), Lido exhibited substantially larger melanopic EDI deviations under specific spectrally narrowband conditions. These large errors were primarily driven by *COLORBOX* conditions, whereas performance under broadband light sources such as *FLUORESCENT* lighting was considerably more stable. This finding highlights the challenges of accurately estimating melanopic quantities under spectrally illumination conditions. The observed device-specific differences are consistent with spectral mismatch effects, whereby sensor spectral responsivities differ from the CIE photopic and melanopic action spectra. Such effects became increasingly pronounced under narrowband and spectrally atypical illumination conditions, where small spectral differences can translate into substantial errors in derived melanopic quantities. *OcuWEAR* demonstrated the greatest variability across all metrics and lighting conditions. Although its performance was acceptable under *SDL* and *Trilux* LED conditions (mean absolute biases ≈ 15–30% for illuminance and melanopic EDI), deviations increased markedly under INCANDESCENT and other LED conditions. For illuminance, mean absolute bias reached 26.9% overall and up to 70% in individual cases (Table S7), while for melanopic EDI, mean absolute bias exceeded 130%, with maximum deviations surpassing 1000%. Under INCANDESCENT lighting alone, *OcuWEAR* exhibited ∼50% error in illuminance and >100% in melanopic EDI, while for CCT, deviations reached up to 155% (Table S8). These results indicate a strong sensitivity to spectral composition, particularly under warm light sources and conditions with significant spectral shifts. Overall, the findings demonstrate that device performance is dependent on both illuminance level and spectral characteristics. While *ActLumus* and *Lido* provided stable and reliable measurements for illuminance across a wide range of lighting conditions, melanopic EDI and CCT remained more challenging metrics, particularly under narrowband or spectrally atypical illumination conditions.

On the other hand, the four light loggers exhibited a high degree of agreement in temporal trends of daylight across all three metrics (illuminance, melanopic EDI and CCT), with differences becoming more pronounced during rapid illumination changes and at values over 10,000 lx m-EDI, possibly due to differing integration periods between devices. The accuracy of wearable light loggers at high melanopic EDI intensities of daylight is likely to be less relevant in field studies, as the ipRGCs and hence the circadian system are likely to be fully saturated above 1000-2000 lx melanopic EDI ^29^. Conversely, the small biases that we found for certain electric light sources under dim light conditions can still be very meaningful. According to Brown et al. ^24^, 10 lx m-EDI is recommended as a cut-off for evening light intensity (starting at least 3 hours before bedtime) measured at the eye in the vertical plane. If this threshold is strictly used in field studies applying wearable light logger devices, any slight under- or overestimation of the device becomes meaningful. Consequently, a device bias of only a few lx m-EDI may result in a different classification of an exposure as compliant or non-compliant with current evening-light recommendations, despite representing a relatively small absolute measurement error. Therefore, based on our results, it seems justified to take the here-reported biases at dim light conditions (< 20 lx m-EDI) into account when reporting the time spent below or above 10 lx m-EDI in the evening hours or the frequency of time intervals above this threshold. Across the tested dim lighting conditions (≤10 lx m-EDI reference levels), absolute bias ranged from approximately 2 to 15 lx m-EDI across devices and light sources (Tables 2, S2-S5). *ActLumus* showed comparatively small deviations (2–4 lx), whereas *Lido* and *OcuWEAR* exhibited larger biases under fluorescent light conditions (up to 9 lx and 15 lx, respectively). Under the lowest tested LED condition (Trilux, ∼46 photopic lx), *OcuWEAR* bias reached 23 lx m-EDI. Given the above-mentioned recommended evening threshold of 10 lx m-EDI, these deviations correspond to more than 100% of the relevant suggested threshold value. Consequently, even moderate systematic over- or underestimation at dim light levels may meaningfully influence threshold-based exposure classification in everyday light exposure monitoring studies. Similar concerns may apply to accurately detecting increased melanopic EDI levels exceeding recommended levels during sleep (≤1 lx m-EDI) ^24^. Importantly, the recommendations proposed by Brown et al. (2022) ^24^ are based on the current understanding of the relationship between melanopic light exposure and human circadian physiology, aiming to promote circadian entrainment, daytime alertness, and healthy sleep while minimising evening and nighttime circadian disruption. Although these recommendations should not be interpreted as strict biological thresholds, they provide a practical framework for evaluating light exposure in research and clinical applications. Consequently, even modest measurement errors near these recommended levels may lead to different classifications of exposure and influence comparisons between studies or the interpretation of compliance with recommended lighting practices.

Regarding the accuracy of wearable light loggers to detect melanopic EDI levels around the current daytime recommendation threshold (≥250 lx m-EDI; ^24^), we found that measurement biases were proportionally smaller compared to those observed under dim evening conditions. Across *FLUORESCENT*, *INCANDESCENT*, and LED conditions at approximately 100–300 lx m-EDI, absolute bias typically ranged between 13 and 47 lx m-EDI for *ActLumus* and between 17 and 44 lx m-EDI for *OcuWEAR* under most light sources, corresponding to roughly 5–20% of the recommended 250 lx m-EDI threshold. *Lido* exhibited larger deviations under *FLUORESCENT* light conditions (up to 173 lx m-EDI, corresponding to approximately 69% of the threshold), and *OcuWEAR* showed increased bias under specific LED configurations (e.g., 128 lx m-EDI under *SDL*, corresponding to approximately 51%). Nevertheless, even in these cases, the relative deviation remained smaller than the bias observed in relation to the 10-lx m-EDI evening threshold. Consequently, classification of exposure above or below the recommended daytime melanopic EDI levels might be more robust to device-specific bias than threshold-based classification under dim evening and night conditions. However, further field-based research is needed to validate these recommendations under everyday light exposure conditions and to determine the extent of absolute and relative measurement bias that can be tolerated without compromising the reliability of exposure classification.

Under dynamic daylight conditions, both *Lido* and *OcuWEAR* exhibited predominantly negative CCT bias relative to the reference spectroradiometer, indicating a systematic underestimation of CCT in daylight. The magnitude of underestimation varied across the daylight spectrum. In the mid-range typical of clear midday conditions (approximately 6500– 8000 K), *Lido* underestimated by roughly 300–500 K (∼4–8% relative deviation), whereas *OcuWEAR* underestimated by approximately 800–1200 K (∼10–18%). At higher CCT levels (>8500 K), corresponding to blue-rich skylight conditions, underestimation became more pronounced, reaching up to −1000 K for *Lido* and up to −3200 K for *OcuWEAR*. In addition, limits of agreement widened substantially at spectral extremes, particularly for *OcuWEAR*, suggesting reduced stability under highly dynamic daylight conditions. From an architectural perspective, such systematic underestimation effectively compresses the dynamic CCT range of daylight, potentially leading to high-CCT daylight conditions being misclassified as closer to standard “daylight white” (≈6500 K). While CCT alone is not a direct predictor of circadian effectiveness, it remains a widely used descriptor in architectural daylight assessment and lighting design practice. Consequently, device-specific CCT bias may influence the evaluation of daylight spectral characteristics in field studies or post-occupancy analyses, particularly when distinguishing between neutral, daylight, and blue-enriched conditions.

While Ishihara et al. recently tested the performance of similar wearable light loggers for measuring photopic and melanopic illuminance under controlled laboratory conditions and naturalistic field settings ^30^, including characterisation of detection ranges, accuracy, and inter-device variability, the present study extends this work by providing more detailed and condition-specific analyses of light logger performance. We further examined how device-specific biases vary across different light sources and measurement contexts, and how these differences influence the interpretation of light exposure metrics. By linking laboratory-based deviations to structured experimental conditions, this study offers additional insight into the mechanisms underlying discrepancies between devices. The only device tested in both their and our present study was the *ActLumus*. In agreement with our findings, Ishihara et al. also found very low inter-device variability as well as highest accuracy across measurement trials (percentage difference to reference spectrometer: +3.3 ± 13.4%, mean ± SD) for the *ActLumus* compared to other devices.

A key factor underlying the observed device- and source-dependent differences is the spectral mismatch inherent in photometric measurements. Most photometric sensors are calibrated using a reference source corresponding to CIE Standard Illuminant A, as defined in international standards (e.g., ISO/CIE 23539:2023; ISO/CIE 11664-2:2022). While this approach ensures comparability and practicality, it introduces systematic deviations when the spectral power distribution of the measured light differs from that of the calibration source. This point is particularly relevant for FLUORESCENT lamps, which exhibit pronounced narrow-band spectral peaks, as well as for LED sources and daylight, whose spectra deviate substantially from tungsten-based Illuminant A. Consequently, similar underlying sensor architectures do not necessarily translate into comparable measurement performance. Although *ActLumus*, *Lido*, and *OcuWEAR* all employ the same 10-channel spectral sensing technology designed to estimate CIE S 026-2018-compliant α-opic light exposure metrics, differences in calibration procedures, spectral weighting functions, and applied mismatch correction factors can result in marked performance differences (CIE ILV, entry 17-25-115). These constraints provide a plausible explanation for the variability observed across devices in the present study, particularly under LED and FLUORESCENT conditions.

Several limitations should be considered when interpreting the findings of the present study. First, the tested light sources may not fully represent the spectral variability encountered in everyday environments. For example, only a single fluorescent spectrum was included, despite substantial variation in spectral power distributions across fluorescent lamp types. Moreover, although the study evaluated a broad range of individually controlled lighting conditions, everyday environments frequently involve mixtures of daylight and electric lighting with dynamically changing spectral compositions. The observed biases may therefore not capture the full range of measurement deviations encountered under everyday living conditions. Future validation studies should include a broader range of spectra and dynamic mixed-light environments to further establish the generalizability of wearable light logger performance. At the same time, the controlled experimental setup represents a strength of the present study, as it enabled systematic comparison of device performance under well-defined conditions.

Second, all measurements were referenced to a single spectroradiometer, which was treated as the criterion instrument. Although spectroradiometers are generally considered highly accurate, residual uncertainty in the reference measurements may have propagated into the reported biases. In addition, the present study was not intended to provide an overall ranking of the evaluated commercial devices, but rather to characterize measurement performance across different lighting conditions and illustrate broader challenges associated with wearable light sensing. Device selection should therefore not be based on measurement accuracy alone; practical considerations such as wearability, user comfort, cost, battery life, and data accessibility may be equally relevant depending on the intended application and research question.

Finally, the potential misclassification of measurements relative to the recommendations proposed by Brown et al. (2022) should be interpreted with caution. These expert consensus-based recommendations provide practical targets for light exposure rather than strict biological thresholds, and circadian and alerting responses to light can vary substantially between individuals and exposure conditions ^31^. Consequently, crossing a recommended value due to measurement uncertainty does not necessarily imply a meaningful difference in the physiological response. Nevertheless, such uncertainty can affect whether an exposure is classified as meeting current recommendations. Recommended melanopic light exposure levels should therefore be interpreted in conjunction with the measurement uncertainty associated with the wearable device and the lighting environment in which measurements are obtained.

In conclusion, our findings demonstrate that wearable light loggers differ substantially in their ability to quantify physiologically relevant light exposure metrics, with agreement relative to the reference spectroradiometer systematically deteriorating as light intensity increased under both electric and daylight conditions. The greatest divergence was observed under high-intensity daylight, suggesting that bright broadband spectra remain particularly challenging for wearable-based estimation of illuminance, CCT, and melanopic EDI. In contrast, dim lighting conditions did not consistently produce the largest absolute disagreement but frequently introduced threshold-relevant bias that may become critical when interpreting light exposure relative to emerging circadian lighting recommendations. Mid-range electric lighting conditions, corresponding approximately to typical indoor daytime environments, yielded comparatively stable performance for the *ActLumus* device, whereas other devices showed less consistent agreement across the same range. Collectively, these findings indicate that wearable device performance is both intensity- and spectrum-dependent, supporting previous observations from field monitoring studies showing that device-specific measurement characteristics can substantially influence estimates of personal light exposure ^19,20,23^. Importantly, by benchmarking wearable devices directly against a research-grade spectroradiometer, the present study provides an empirical framework for interpreting wearable-derived light exposure data within circadian, sleep, and health research contexts, where even relatively small measurement inaccuracies may alter inferred relationships between light exposure and biological outcomes. These results are also relevant for architectural lighting research and evidence-based lighting design, where wearable assessments are increasingly used to evaluate the circadian effectiveness of built environments. In this context, appropriate device selection and awareness of condition-specific measurement limitations are essential to ensure that occupant-level melanopic and photopic exposure estimates accurately reflect the lighting conditions experienced in everyday environments.

## Methods

### Experimental set-ups

To evaluate agreement across representative wearable measurement technologies currently used in circadian and sleep research, three wearable light loggers (*ActLumus*, Condor Instruments, São Paulo, Brazil; *Lido*, Lucerne School of Engineering and Architecture, Horw, Switzerland; and *OcuWEAR 1.0*, Ocutune, Roskilde, Denmark) were selected and cross-validated against an advanced spectroradiometer (*CSS-45*, Gigahertz Optik GmbH, Türkenfeld, Germany). The three wearable light loggers were selected because they represent contemporary research-grade devices currently used for personal light exposure assessment in circadian and sleep research and were available through the collaborating research groups. Their differing hardware designs and sensing approaches enabled comparison across a range of wearable measurement technologies. All devices were placed in a custom-designed styrofoam testbed next to each other horizontally (Figure 1) and configured to sample at 0.1 Hz. We tested several different lighting conditions: FLUORESCENT ceiling light in a laboratory office room with a ramp protocol from minimum to maximum light intensities (≈10-2,000 photopic lx), LED-COLORBOX of different colours set to a constant intensity of about 100 photopic lx (green and red), DAYLIGHT with an open window from an office room in Aachen (Germany) on one winter day with varying daylight intensities (≈2,000-30,000 photopic lx), different typical light bulbs from minimum to maximum light intensities (INCANDESCENT at 2,800 K from ≈10-600 photopic lx, LED at 4,000 K from ≈40-4,400 photopic lx) and a daylight-mimicking LED standing lamp (≈5,800 K from ≈100-13,000 photopic lx). Every different indoor light condition or respective intensity was tested for 5 minutes twice. All indoor experiments were conducted at typical room temperature (21-24°C, 30-50% humidity). To test different colour settings of LED lighting and fluorescent light at different intensities, we utilised the technical light exposure laboratory at the Eindhoven University of Technology (TU/e). To further test other typical indoor light conditions, we built a customised cardboard box covered with white glossy photo paper, in which several electric light sources, described in detail below, were installed together with the wearable light loggers and the reference spectroradiometer.

### Enclosed measurement setup

For several electric light conditions (INCANDESCENT and selected LED configurations), a custom-made cardboard enclosure (Figure 2A-B) was used. Although the enclosure does not represent a standardised photometric integrating environment, it enabled controlled and repeatable comparisons of device responses under different electric light sources. Different electric light sources, including light bulbs and standing lamps, were installed inside the enclosure together with the wearable light loggers and the reference spectroradiometer. All devices were mounted in the same custom-designed styrofoam testbed described above to ensure identical measurement geometry. The approximate distance between the light loggers and the respective light source within the cardboard box was 30-40 cm, depending on the physical dimension of the light source (Figure 2B). During these experiments, the *Lido* device was no longer available and was therefore excluded from these electric lighting measurements. This enclosure was used for the INCANDESCENT, SDL, Trilux, and tunable LED (Vitae) conditions described below.

### FLUORESCENT light

To assess performance under typical office lighting conditions, a FLUORESCENT lighting setup was used. A laboratory set-up of fluorescent tube lamps installed in a grid ceiling was used (DALI controllable Philips Savio ceiling fixtures). The wearable light loggers and the reference spectroradiometer were oriented horizontally and directed toward a vertical wall, simulating frontal light exposure of an office worker sitting at a desk (Figure 11B).

A stepwise light intensity ramp from 10 (minimum) to 2000 (maximum) photopic lx (10, 100, 300, 1000, and 2000 lx) at an average CCT of 4800 K as measured by the reference spectroradiometer, was applied twice.

### INCANDESCENT light bulb

To evaluate performance under warm, broadband spectra, incandescent lighting conditions were tested. We installed an incandescent light bulb (XQ Lite, E14, 28 W, 2800-3000 K) within the cardboard enclosure (Figure 2C). A stepwise illuminance ramp was applied, comprising target photopic illuminance levels of 10 lx (minimum), 100, 300 to 600 photopic lx (maximum), as measured by the reference spectroradiometer. Between repetitions, the light source and sensor configuration remained unchanged.

### LED conditions

To systematically assess device performance under LED-based lighting, several configurations were tested, varying in spectral composition and intensity:

#### COLORBOX

This setup enabled controlled testing under narrowband spectral conditions. To test green and red colours of LED light, the custom-engineered Boschman ColorBox was used (Boschman 2021). It contained 11 LED types, each with a different spectrum and individually controlled, to produce a Ganzfeld-like light exposure (Figure 10A). In these experiments, an additional handheld spectrometer (*MK350D*, UPRtek, Zhunan, Taiwan) was applied next to the three wearable light loggers and the reference spectroradiometer, but due to technical issues, its recorded data were eventually not analysed.

**Figure 10:**
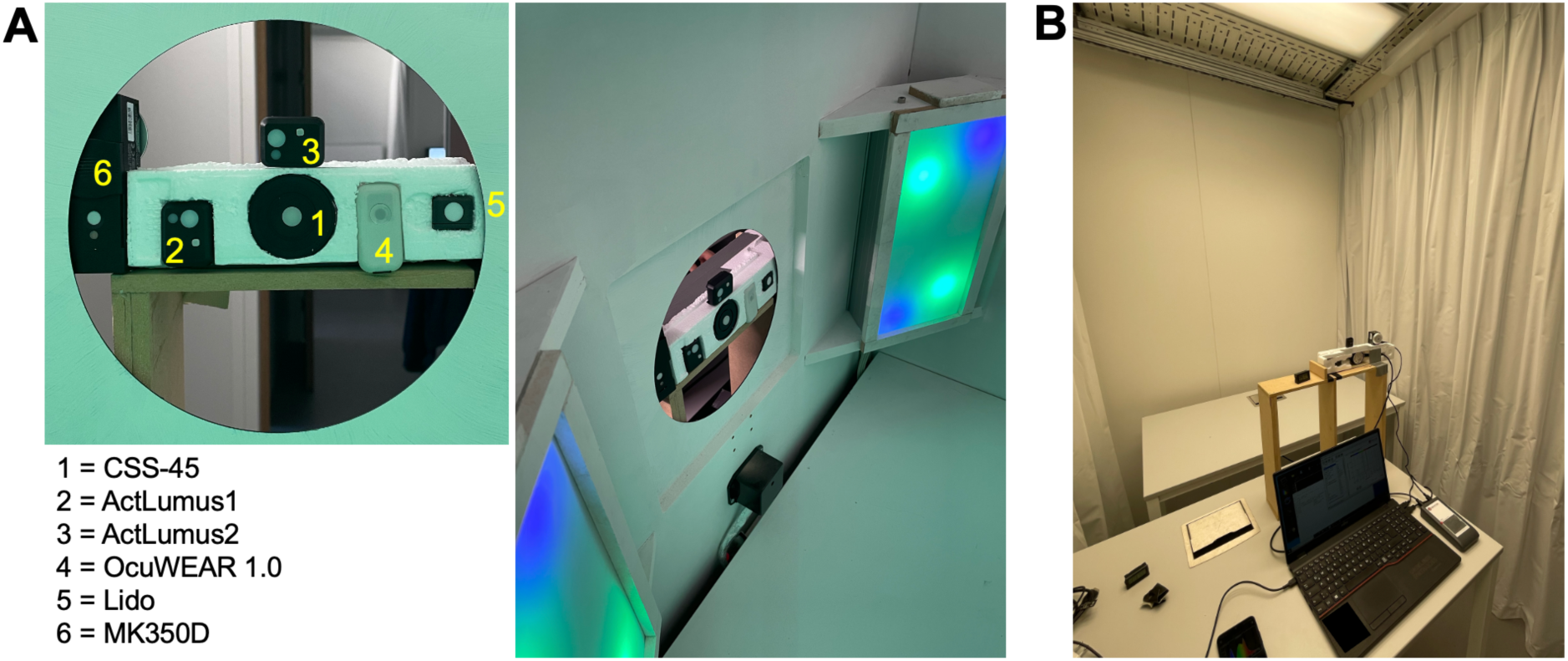
Experimental set-up at TU/e for the LED-ColorBox (A) and FLUORESCENT light condition (B).

##### Standing DAYLIGHT-MIMICKING LED lamp (SDL)

A daylight-mimicking LED standing lamp (Spectrasol Terra, Spectrasol s.r.o., Prague, Czech Republic) was positioned horizontally, with the downward-facing emitting surface directed to the measurement set-up. To maintain a consistent enclosure geometry, the cardboard box was also rotated accordingly and closed with white photographic paper (Figure 2G). This configuration was selected to expose the devices to a wide range of high-intensity electric light while maintaining a controlled and repeatable measurement geometry. A stepwise illuminance ramp was applied, comprising target photopic illuminance levels of approximately 130 lx (minimum), 300 lx to 13,000 lx (maximum) as measured by the reference spectroradiometer.

##### LED standing lamp (Trilux)

Another LED standing lamp (LuceoS Act S, TRILUX GmbH, Arnsberg, Germany) was positioned horizontally, with the downward-facing light directed towards the measurement set-up, while the cardboard enclosure was rotated accordingly (Figure 2H). The enclosure was subsequently closed using white photographic paper, following the same configuration as applied for the daylight-mimicking LED lamp. We set the LED standing lamp to a constant CCT of 4,000 K. The upward-emitting light component was set to its minimum intensity of 1% using a smartphone-based control application (LiveLink Workplace, TRILUX GmbH), while only the downward-emitting light was manipulated during the experiment. A stepwise illuminance ramp was applied, comprising approximate target photopic illuminance levels of 50 lx (minimum), 300 lx and 4300 lx (maximum), as measured by the reference spectroradiometer. These levels corresponded to programmed lamp intensities of 1%, 19% and 100%, respectively.

##### TUNABLE LED light bulb (Vitae)

We installed a commercially available LED light bulb (Lumie Vitae, E14, 28 W, Prague, Czech Republic) within the cardboard enclosure. This light source features three discrete manufacturer-defined CCT settings called *Orange* to use at night (unspecified Kelvin by manufacturer; Figure 2D), *Warm White* to use in the evening after dusk (2700 K; Figure 2E), and *White* to use during the solar day (4000 K, Figure 2F). Due to technical limitations of the experimental setup, dimming of the LED light sources was not possible. Therefore, we did not conduct a light intensity ramp protocol. Instead, each CCT setting was measured at its default operating intensity after switch-on. Measurements were conducted once per setting. Photopic illuminance levels recorded by the reference spectroradiometer were approximately 60 lx (*Orange*), 1500 lx (*Warm White*), and 2000 lx (*White*).

To facilitate interpretation of the experimental protocol, the planned lighting conditions together with the corresponding reference measurements are summarised in Table 2. For each lighting condition, the table reports the number of repeated measurements and the reference photopic illuminance, melanopic EDI, and CCT measured using the CSS-45 spectroradiometer. These reference values are presented as mean ± standard deviation and represent the actual lighting conditions included in the agreement analyses.

**Table 2:**
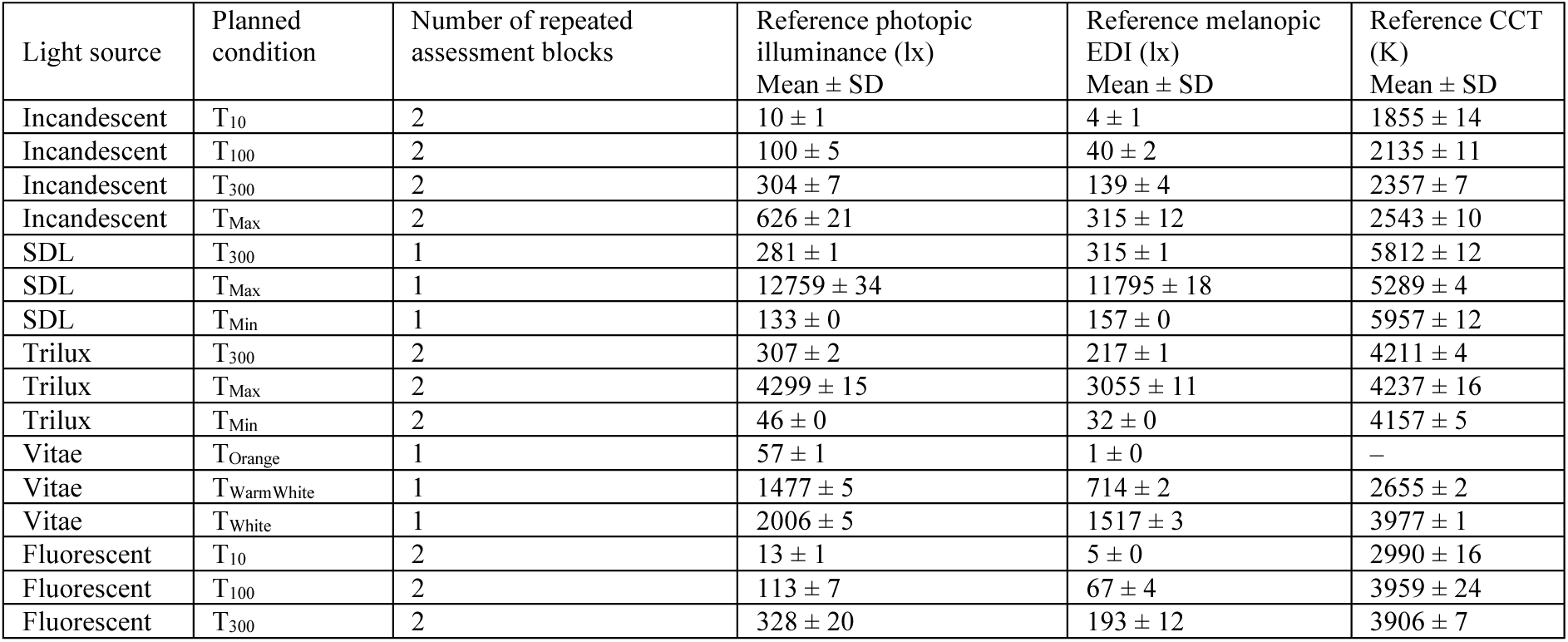
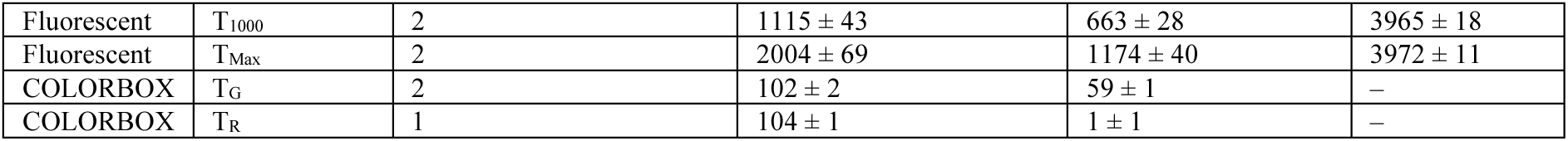
Summary of the laboratory lighting conditions, including the planned lighting condition, the number of repeated measurements, and the corresponding reference photopic illuminance, melanopic EDI, and CCT measured using the CSS-45 spectroradiometer. Reference values are reported as mean ± standard deviation (SD). Most laboratory lighting conditions were repeated twice to assess measurement repeatability. Conditions evaluated only once are indicated accordingly.

### DAYLIGHT

To complement controlled laboratory measurements, device performance was also evaluated under dynamic natural daylight conditions. Daylight measurements were conducted in a suburb of Aachen, Germany (50.8°N, 6.0°E). The three wearable light loggers and the reference spectroradiometer were mounted together in the same custom-designed styrofoam testbed (Figure 1) to ensure identical measurement geometry. The testbed was placed immediately inside the façade line of an office room, with the window open to avoid spectral attenuation by glazing (Figure 11A). Measurements were performed on January 9, 2025, from 09:00 to 15:00 CET, coinciding with local sunrise in Aachen (08:35 CET). Weather conditions included a snowy ground cover and generally clear morning conditions with intermittent cloud cover. During the measurement period, photopic illuminance was recorded by the reference spectrometer, ranging from approximately 2,000 lx to 30,000 lx, capturing a broad range of high-intensity daylight conditions (Figure 11B).

**Figure 11:**
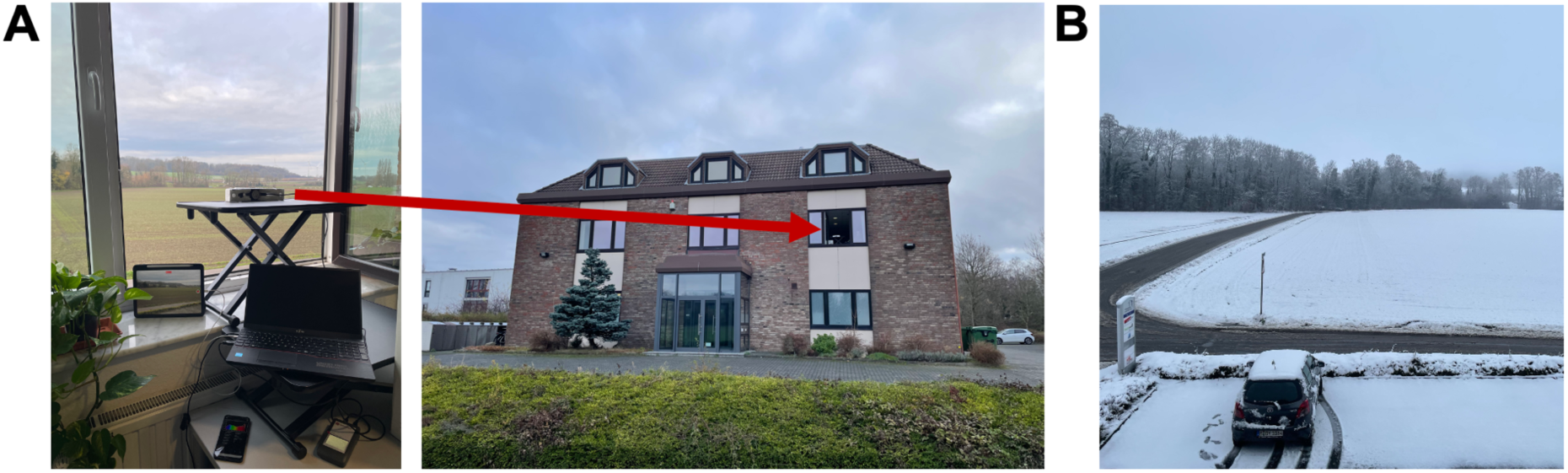
Experimental set-up to test dynamic natural daylight conditions (A) on a snowy winter day (B).

### Data analyses and statistics

The following analysis approach was applied consistently across all light conditions described above. Raw photopic illuminance, melanopic EDI, and CCT data were collected from all light loggers and next processed in R (v4.3.1). Before data matching, device-specific clock offsets were corrected using the recorded timestamps and visual inspection of the synchronised time series. The CSS-45 measurements served as the temporal reference. For each CSS-45 observation, the temporally closest measurement from each wearable light logger was identified using a nearest-neighbour matching algorithm (k = 1), implemented using the get.knnx function from the FNN package ^32^. No interpolation or temporal averaging was performed before matching; paired observations were generated directly from the nearest available timestamps. The resulting paired measurements for photopic illuminance, melanopic EDI, and CCT were stored in a merged dataset for subsequent analyses.

Because each lighting condition remained stable for five minutes and measurements acquired during lighting transitions were excluded from the analyses, a 10-s sampling interval yielded approximately 30 paired observations per condition, which was considered appropriate to characterise device agreement under the stable lighting conditions investigated in this study. For the Lido, the sensor integration time is automatically adjusted according to incident light level and ranges from 2.8 to 716.8 ms per conversion (manufacturer communication), remaining below the 10-s sampling interval even under dim-light conditions.

For each 5-minute light condition, agreement between each wearable light logger and the reference spectroradiometer was assessed using Bland–Altman analysis ^33^. Bias (mean signed difference) was calculated as the arithmetic mean of the paired differences between the wearable light logger and the reference spectroradiometer. Positive values indicate systematic overestimation relative to the reference, whereas negative values indicate systematic underestimation. Accordingly, the differences were intentionally not converted to absolute values, as preserving the sign is fundamental to Bland–Altman analysis. Formally, bias was defined as:

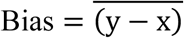

where x denotes the reference measurements obtained with the *CSS-45*, and y denotes measurements from the wearable light loggers. The 95% Limits of Agreement (LoA) were calculated to quantify the range within which 95% of paired measurement differences are expected to lie, capturing both systematic and random measurement error. The limits were defined as:

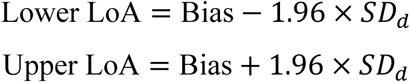

where *SD_d_* is the standard deviation of the paired differences.

The factor 1.96 corresponds to the approximate 95% coverage expected when the paired measurement differences are normally distributed, as assumed in the classical Bland–Altman method. To facilitate comparison across lighting conditions with substantially different absolute levels, bias and limits of agreement were additionally normalised to the mean *CSS-45* value of each scenario. This procedure yielded relative agreement metrics that are independent of absolute light metrics:

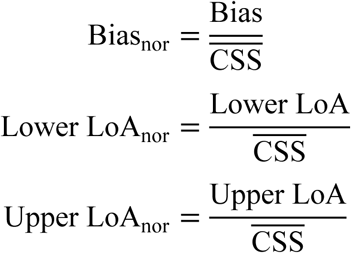

where 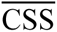 denotes the mean *CSS-45* reference value within the corresponding scenario. Agreements were evaluated using Bland-Altman statistics implemented via the *blandr.statistics* function from the blandr package ^34^. Missing paired observations occurred in some scenarios due to intermittent data recording by certain wearable devices (e.g., temporary logging interruptions or synchronisation mismatches). Lighting conditions with fewer than five valid paired observations were excluded from the Bland-Altman analysis. Time windows affected by sudden lighting transitions were manually cleaned by assigning missing values to affected data points. Aggregating bias and limits of agreement by light source, light logger, and metric enabled the identification of systematic performance patterns across lighting conditions, while grouped ribbon plots provided a compact visual representation of both central bias and variability, highlighting source-dependent differences in light logger agreement.

In addition, agreement between two *ActLumus* configurations close to each other (Figure 10A) was assessed across valid *COLORBOX* and *FLUORESCENT* scenarios. For each matched pair of illuminances and melanopic EDI values, scatterplots were generated, and linear regressions were fitted using least-squares estimation. Coefficients of determination (R²) were extracted to quantify inter-device consistency and assess interchangeability or systematic deviations between configurations.

A separate analysis was conducted to evaluate the performance of wearable light loggers under dynamic natural daylight conditions. Unlike the artificial lighting experiments, the daylight dataset was not segmented into predefined light conditions. This decision was intentional, as the primary objective was to preserve the continuous temporal dynamics of everyday daylight exposure rather than to assess light logger agreements under controlled, quasi-steady states. Time-series plots were generated for light metrics to visualise the evolution of daylight exposure across the measurement period. Due to the wide dynamic range of daylight, spanning several orders of magnitude, a base-10 logarithmic scaling was applied to the y-axes of the photopic illuminance and melanopic EDI time-series plots to improve interpretability and prevent high-intensity periods from visually dominating the lower-intensity measurements. Similar to electric lighting analysis and to ensure comparability between devices with differing internal clocks, all wearable logger measurements were temporally aligned to the reference spectroradiometric system. Then, for each CSS timestamp, the nearest measurement in each wearable device dataset was identified. To assess agreement between measurement methods under continuously changing daylight conditions, we adapted the traditional Bland–Altman method. Classical Bland–Altman analysis assumes constant bias, meaning that the average difference between measurements is independent of measurement magnitude, and homoscedastic residuals, meaning that the variability of the differences remains constant across the measurement range ^35^. These assumptions are frequently violated under highly dynamic daylight conditions, where both measurement bias and variance may systematically vary with light level, motivating the complementary LOESS-based analysis presented here. To account for these violations, we applied locally weighted scatterplot smoothing (LOESS) ^36^ to the Bland-Altman residuals. LOESS is a non-parametric smoothing technique that fits simple local regressions within small neighbourhoods of the data and joins them smoothly, without assuming a predefined global functional form. In contrast to a single global regression line, LOESS allows the relationship between measurement differences and magnitude to vary gradually across the measurement range.

Applying LOESS to the Bland–Altman residuals enables a visual assessment of magnitude-dependent bias, nonlinearity, and heteroscedasticity ^37^, which are common under high dynamic range lighting conditions such as daylight exposure. Specifically, deviations of the LOESS curve from a horizontal line indicate non-constant bias, while curved patterns reveal systematic nonlinear effects that are not captured by the mean difference alone. This approach may also uncover structured trends that are obscured by the classical assumption of constant bias. For each light metric and measurement pair, the mean of the paired *CSS-45* and wearable light logger measurements was calculated, and the difference between the wearable light logger and *CSS-45* reference was computed. A LOESS curve was then fitted to the residuals as a function of the mean values (smoothing span = 0.75), and smoothed residual plots were generated to visualise how measurement bias evolved across the full range of light levels. This approach mitigates the assumption of constant bias inherent in traditional Bland-Altman analysis and enables local estimation of agreement across different intensity ranges. As a result, the visualisations provide a more nuanced representation of measurement agreement across the range of daylight conditions encountered in everyday life and support the identification of light-level-dependent deviations that may not be evident from summary statistics alone.

In the LOESS-based Bland–Altman plots, custom x- and y-axis scales were applied using pseudo-logarithmic transformations (*pseudo_log_trans* function from scales package; Wickham et al., 2025) to accommodate the wide dynamic range of daylight measurement values while maintaining interpretability near zero. This scaling is particularly appropriate to daylight exposure, which varied over several orders of magnitude during the measurement period. To align with visualisations with known properties of human visual sensitivity and psychophysiological relevance, x-axis breaks for photopic illuminance and melanopic EDI were selected based on established perceptual thresholds, including values representative of scotopic conditions, typical indoor lighting, bright daylight, and direct sunlight (CIE 117-1995 / Daylight; ^38^. For CCT, axis breaks at 2700 K, 4000 K, 6500 K, and 10,000 K were used to capture perceptually meaningful categories corresponding to warm white, neutral white, daylight, and high-CCT blue-enhanced light consistent with standardised white-point classifications (CIE 248:2022 – Colour appearance models). This scaling strategy reflects the non-linear nature of human light perception, consistent with classical psychophysical principles described by the Weber–Fechner law ^39^ and Stevens’ Power Law ^40^. At high illuminance levels, such as daylight exceeding 10,000 lx, the system exhibits reduced sensitivity to small absolute changes, rendering modest increases largely imperceptible. Similarly, melanopic stimulation shows a saturating response, with ipRGC-mediated circadian activation beginning to plateau at approximately 1,000–2,000 lx m-EDI, beyond which additional light produces diminishing physiological effects (CIE S 026/E:2018 – System for Metrology of Optical Radiation for ipRGC-Influenced Responses to Light; ^29^. By applying perceptually informed pseudo-log scaling, the resulting plots highlight variations that are most relevant to visual comfort and circadian regulation, while avoiding distortion caused by over-compressing of low-value ranges or over-emphasising extreme values.

## Data availability

The datasets generated and analysed during the current study are not publicly available because they form part of ongoing and follow-up research, but are available from the corresponding author on reasonable request.

## Code availability

The underlying code used for data processing and statistical analysis is not publicly available but may be made available to qualified researchers on reasonable request from the corresponding author.

## Acknowledgements

Peiman Pilehchi-Ha and Marcel Schweiker were supported by a research fund (21055) by VILLUM FONDEN. Jan-Frieder Harmsen is funded by the project “Profilbildung Built and Lived Environment” (PB22-062). The project “Profilbildung Built and Lived Environment” has received funding from the programme “Profilbildung 2022”, an initiative of the Ministry of Culture and Science of the State of North Rhine-Westphalia. The sole responsibility for the content of this publication lies with the authors. We thank *Ocutune* for providing us with their prototype *OcuWEAR*1.0 for the duration of the study; the company neither influenced the conducted experiments, data analyses, nor drew conclusions. We further thank the staff of the Humans & Technology laboratory of Eindhoven University of Technology for their assistance.

## Author contributions

Conception or design of the work: PPH, OS, and JFH. Acquisition, analysis or interpretation of data for the work: all authors. Drafting the work or revising it critically for important intellectual content: all authors.

## Declaration of generative AI and AI-assisted technologies in the writing process

During the preparation of this work, the authors used ChatGPT to polish the text of the manuscript. After using this tool/service, the authors reviewed and edited the content as needed and take full responsibility for the content of the publication.

## Supplemental Appendix

**Supplemental Figure S1:**
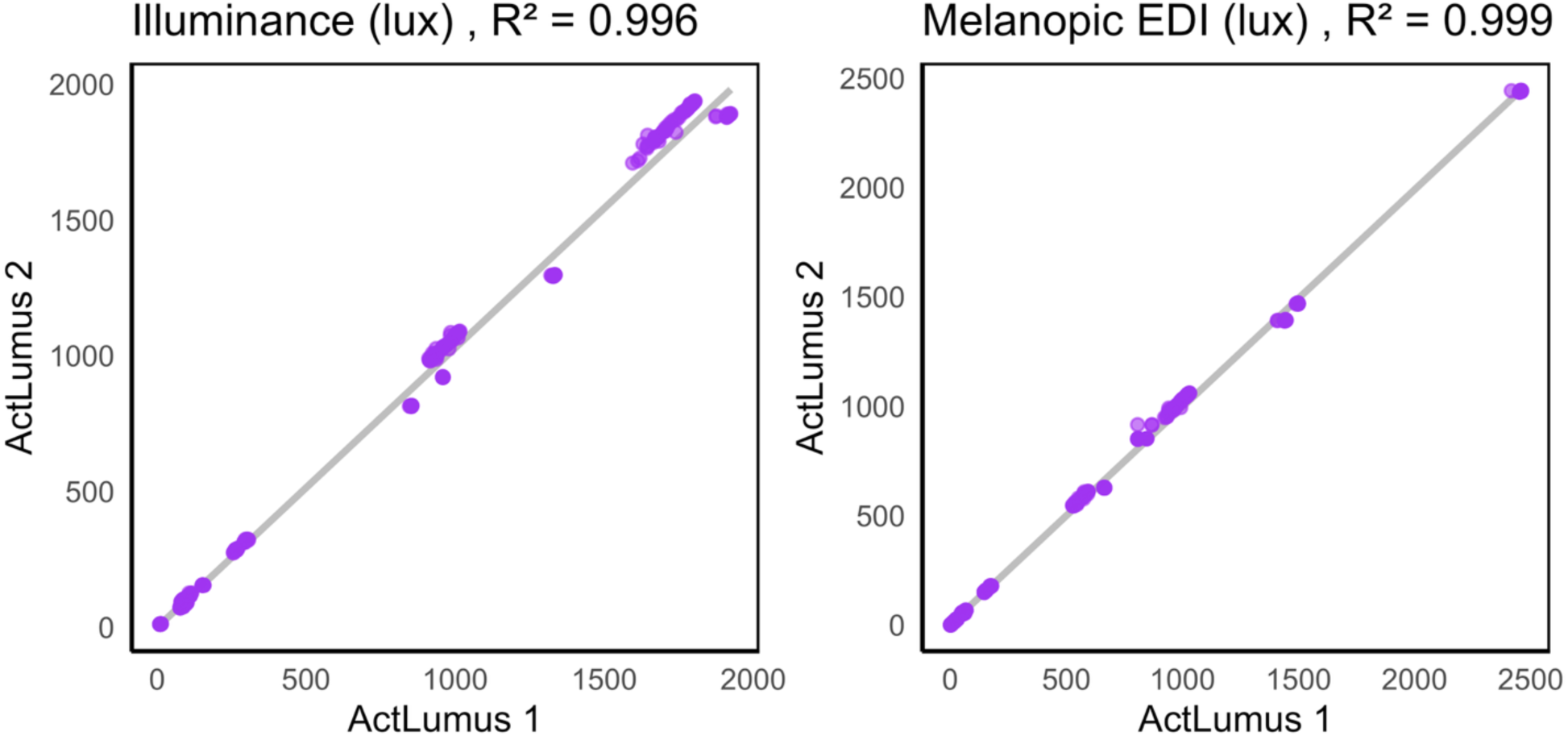
Comparison of the measured illuminance (A) and melanopic EDI (B) values in two ActLumus sensors closely placed next to each other (see Figure 3A) in the COLORBOX and FLUORESCENT experiments.

**Supplemental Table S1.** Bland-Altman results for light loggers exposed to the INCANDESCENT light source. T_max_ was 550 lx measured by CSS. A positive bias value indicates that the respective light logger device displayed higher values compared to the reference spectroradiometer, and vice versa for negative values.

| Light logger | Metric | Target condition | Bias | Lower LoA | Upper LoA |
| --- | --- | --- | --- | --- | --- |
| *OcuWEAR* | Illuminance (lx) | T<sub>10</sub> | -3 | -28 | 22 |
|  |  | T<sub>100</sub> | -70 | -318 | 178 |
|  |  | T<sub>300</sub> | 142 | 132 | 151 |
|  |  | T<sub>Max</sub> | 332 | 304 | 361 |
|  | Melanopic EDI (lx) | T<sub>10</sub> | -8 | -25 | 10 |
|  |  | T<sub>100</sub> | -63 | -189 | 63 |
|  |  | T<sub>300</sub> | 23 | 17 | 29 |
|  |  | T<sub>Max</sub> | 111 | 95 | 128 |
|  | CCT (K) | T<sub>10</sub> | -2871 | -3158 | -2584 |
|  |  | T<sub>100</sub> | -1330 | -2014 | -645 |
|  |  | T<sub>300</sub> | -1127 | -1190 | -1064 |
|  |  | T<sub>Max</sub> | -827 | -883 | -772 |
| *ActLumus* | Illuminance (lx) | T<sub>10</sub> | -2 | -2 | -1 |
|  |  | T<sub>100</sub> | -1 | -3 | 0 |
|  |  | T<sub>300</sub> | 10 | 2 | 18 |
|  |  | T<sub>Max</sub> | 32 | 17 | 46 |
|  | Melanopic EDI (lx) | T<sub>10</sub> | 3 | 3 | 4 |
|  |  | T<sub>100</sub> | 17 | 15 | 18 |
|  |  | T<sub>300</sub> | 43 | 38 | 47 |
|  |  | T<sub>Max</sub> | 79 | 61 | 98 |

**Supplemental Table S2.**
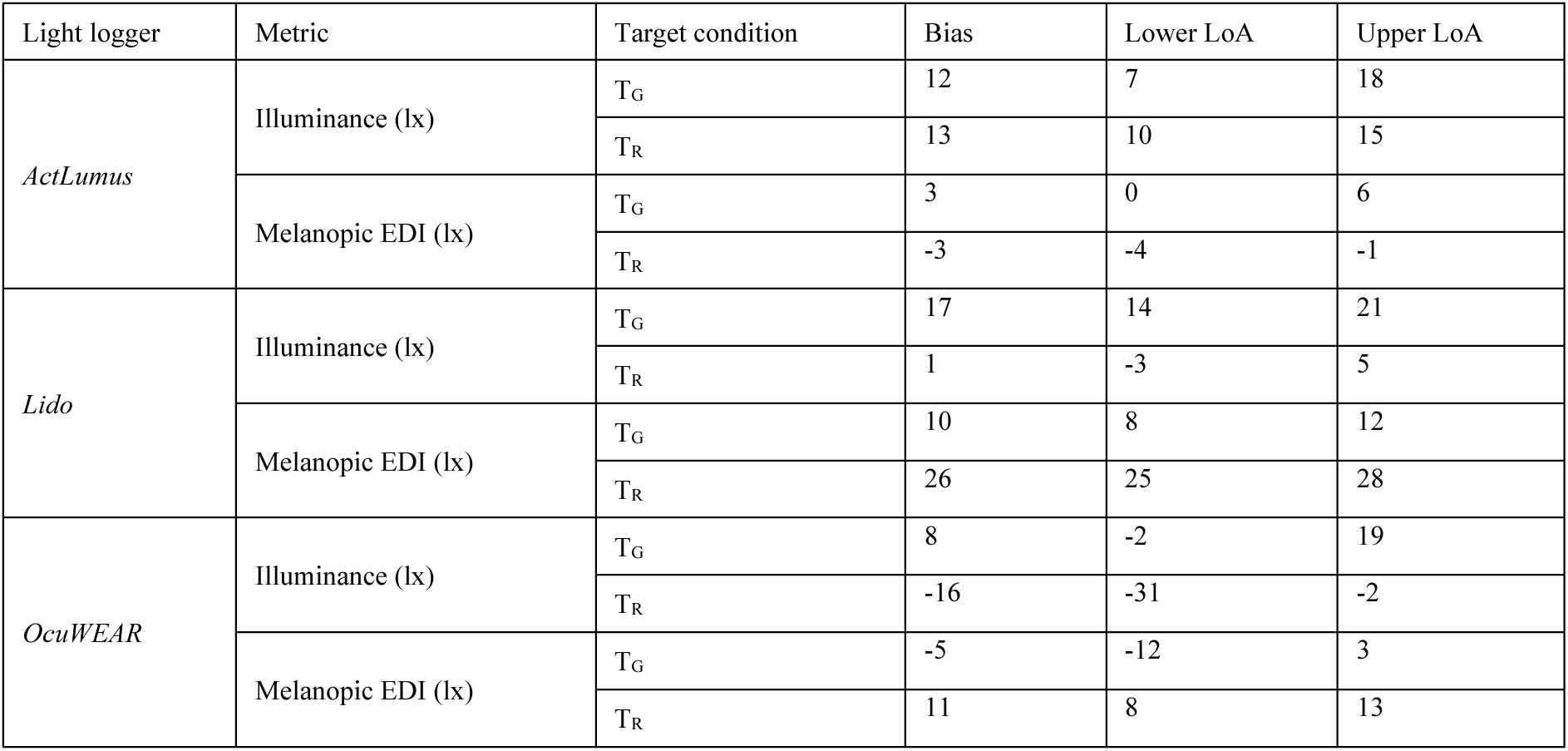
Bland-Altman results for light loggers exposed to COLORBOX light source. T_G_ is Light Green (102 lx) and T_R_ is Red (104 lx). A positive bias value indicates that the respective light logger device displayed higher values compared to the reference spectroradiometer, and vice versa for negative values.

| Light logger | Metric | Target condition | Bias | Lower LoA | Upper LoA |
| --- | --- | --- | --- | --- | --- |
| *ActLumus* | Illuminance (lx) | $T_G$ | 12 | 7 | 18 |
| | | $T_R$ | 13 | 10 | 15 |
| | Melanopic EDI (lx) | $T_G$ | 3 | 0 | 6 |
| | | $T_R$ | -3 | -4 | -1 |
| *Lido* | Illuminance (lx) | $T_G$ | 17 | 14 | 21 |
| | | $T_R$ | 1 | -3 | 5 |
| | Melanopic EDI (lx) | $T_G$ | 10 | 8 | 12 |
| | | $T_R$ | 26 | 25 | 28 |
| *OcuWEAR* | Illuminance (lx) | $T_G$ | 8 | -2 | 19 |
| | | $T_R$ | -16 | -31 | -2 |
| | Melanopic EDI (lx) | $T_G$ | -5 | -12 | 3 |
| | | $T_R$ | 11 | 8 | 13 |

**Supplemental Table S3.** Bland-Altman results for light loggers exposed to SDL. T_Min_ and T_Max_ were 133 and 12759 photopic lx, respectively, measured by CSS. A positive bias value indicates that the respective light logger device displayed higher values compared to the reference spectroradiometer, and vice versa for negative values.

| Light logger | Metric | Target condition | Bias | Lower LoA | Upper LoA |
| --- | --- | --- | --- | --- | --- |
| *ActLumus* | Illuminance (lx) | $T_{Min}$ | 18 | 18 | 19 |
| | | $T_{300}$ | 38 | 37 | 40 |
| | | $T_{Max}$ | 1397 | 1372 | 1421 |
| | Melanopic EDI (lx) | $T_{Min}$ | 24 | 23 | 25 |
| | | $T_{300}$ | 47 | 46 | 49 |
| | | $T_{Max}$ | 1190 | 1170 | 1210 |
| *OcuWEAR* | Illuminance (lx) | $T_{Min}$ | -6 | -7 | -5 |
| | | $T_{300}$ | 63 | 56 | 70 |
| | | $T_{Max}$ | -2456 | -2529 | -2383 |
| | Melanopic EDI (lx) | $T_{Min}$ | 34 | 34 | 35 |
| | | $T_{300}$ | 128 | 123 | 134 |
| | | $T_{Max}$ | -3452 | -3701 | -3203 |
| | CCT (K) | $T_{Min}$ | 1159 | 1129 | 1188 |
| | | $T_{300}$ | 1291 | 1256 | 1326 |
| | | $T_{Max}$ | -679 | -801 | -557 |

**Supplemental Table S4.** Bland-Altman results for light loggers exposed to Trilux Lamp. T_Min_ and T_Max_ were 46 lx and 4299 lx, respectively, measured by CSS. A positive bias value indicates that the respective light logger device displayed higher values compared to the reference spectroradiometer, and vice versa for negative values.

| Light logger | Metric | Target condition | Bias | Lower LoA | Upper LoA |
| --- | --- | --- | --- | --- | --- |
| *ActLumus* | Illuminance (lx) | $T_{Min}$ | 7 | 6 | 7 |
| | | $T_{300}$ | 41 | 37 | 44 |
| | | $T_{Max}$ | 519 | 510 | 528 |
| | Melanopic EDI (lx) | $T_{Min}$ | 4 | 4 | 4 |
| | | $T_{300}$ | 23 | 21 | 26 |
| | | $T_{Max}$ | 281 | 264 | 299 |
| *OcuWEAR* | Illuminance (lx) | $T_{Min}$ | 8 | 8 | 9 |
| | | $T_{300}$ | 17 | 14 | 20 |
| | | $T_{Max}$ | 116 | 106 | 126 |
| | Melanopic EDI (lx) | $T_{Min}$ | 23 | 23 | 23 |
| | | $T_{300}$ | 17 | 15 | 19 |
| | | $T_{Max}$ | -118 | -137 | -99 |
| | CCT (K) | $T_{Min}$ | 1507 | 1496 | 1518 |
| | | $T_{300}$ | 236 | 223 | 248 |
| | | $T_{Max}$ | -143 | -174 | -111 |

**Supplemental Table S5.** Bland-Altman results for ActLumus exposed to Vitae light bulb. T_orange_, T_warmwhite_ and T_white_ were equal to 57 lx, 1477 lx and 2006 lx, respectively, measured by CSS. A positive bias value indicates that the respective light logger device displayed higher values compared to the reference spectroradiometer, and vice versa for negative values.

| Light logger | Metric | Target condition | Bias | Lower LoA | Upper LoA |
| --- | --- | --- | --- | --- | --- |
| *ActLumus* | Illuminance (lx) | $T_{orange}$ | 14 | 14 | 15 |
| | | $T_{warmwhite}$ | 199 | 196 | 202 |
| | | $T_{white}$ | 274 | 271 | 278 |
| | Melanopic EDI (lx) | $T_{orange}$ | 0 | 0 | 1 |
| | | $T_{warmwhite}$ | 91 | 89 | 93 |
| | | $T_{white}$ | 193 | 190 | 196 |
| *OcuWEAR* | Illuminance (lx) | $T_{orange}$ | -9 | -9 | -8 |
| | | $T_{warmwhite}$ | 875 | 868 | 881 |
| | | $T_{white}$ | 1277 | 1015 | 1539 |
| | Melanopic EDI (lx) | $T_{ON}$ | 5 | 5 | 6 |
| | | $T_{WE}$ | 483 | 476 | 490 |
| | | $T_{WD}$ | 1053 | 879 | 1227 |
| | CCT (K) | $T_{ON}$ | NA | NA | NA |
| | | $T_{WE}$ | 258 | 232 | 283 |
| | | $T_{WD}$ | 637 | 140 | 1133 |

**Supplemental Table S6.**
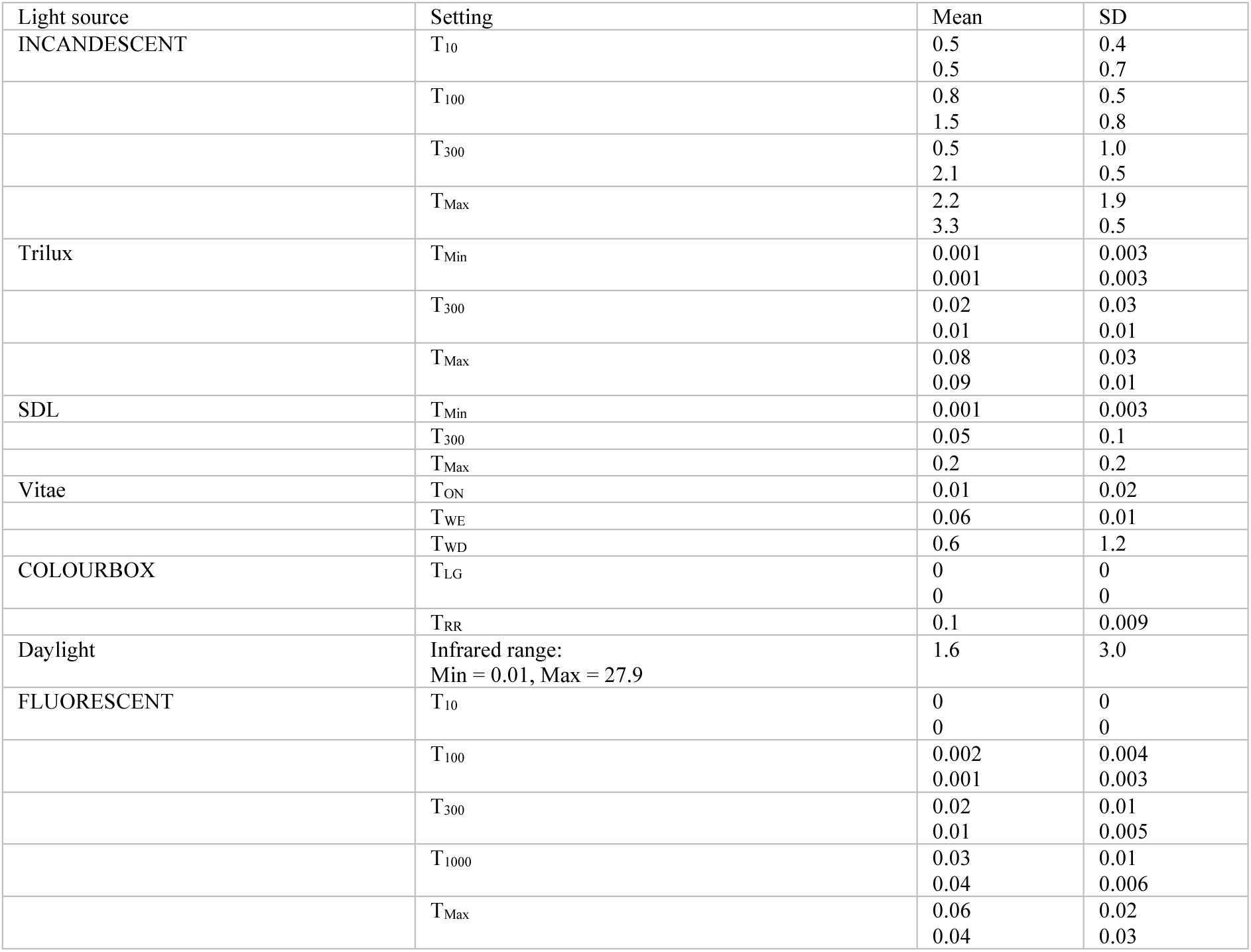
Infrared light quantification (³ 850 nm) in mW/cm^2^ derived from the ActLumus device in all the different light source conditions; mean ± standard deviation (SD) is reported in chronological order of the two 5-min data collection runs.

**Supplemental Table S7.**
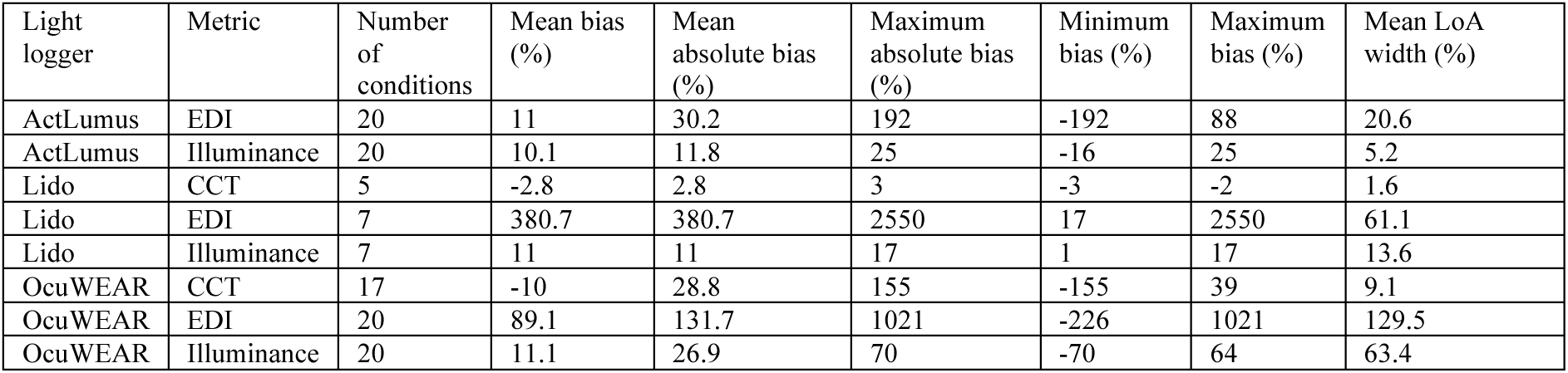
Summary of relative bias and agreement metrics (%) across all lighting conditions and devices

| Light logger | Metric | Number of conditions | Mean bias (%) | Mean absolute bias (%) | Maximum absolute bias (%) | Minimum bias (%) | Maximum bias (%) | Mean LoA width (%) |
| --- | --- | --- | --- | --- | --- | --- | --- | --- |
| ActLumus | EDI | 20 | 11 | 30.2 | 192 | -192 | 88 | 20.6 |
| ActLumus | Illuminance | 20 | 10.1 | 11.8 | 25 | -16 | 25 | 5.2 |
| Lido | CCT | 5 | -2.8 | 2.8 | 3 | -3 | -2 | 1.6 |
| Lido | EDI | 7 | 380.7 | 380.7 | 2550 | 17 | 2550 | 61.1 |
| Lido | Illuminance | 7 | 11 | 11 | 17 | 1 | 17 | 13.6 |
| OcuWEAR | CCT | 17 | -10 | 28.8 | 155 | -155 | 39 | 9.1 |
| OcuWEAR | EDI | 20 | 89.1 | 131.7 | 1021 | -226 | 1021 | 129.5 |
| OcuWEAR | Illuminance | 20 | 11.1 | 26.9 | 70 | -70 | 64 | 63.4 |

**Supplemental Table S8:**
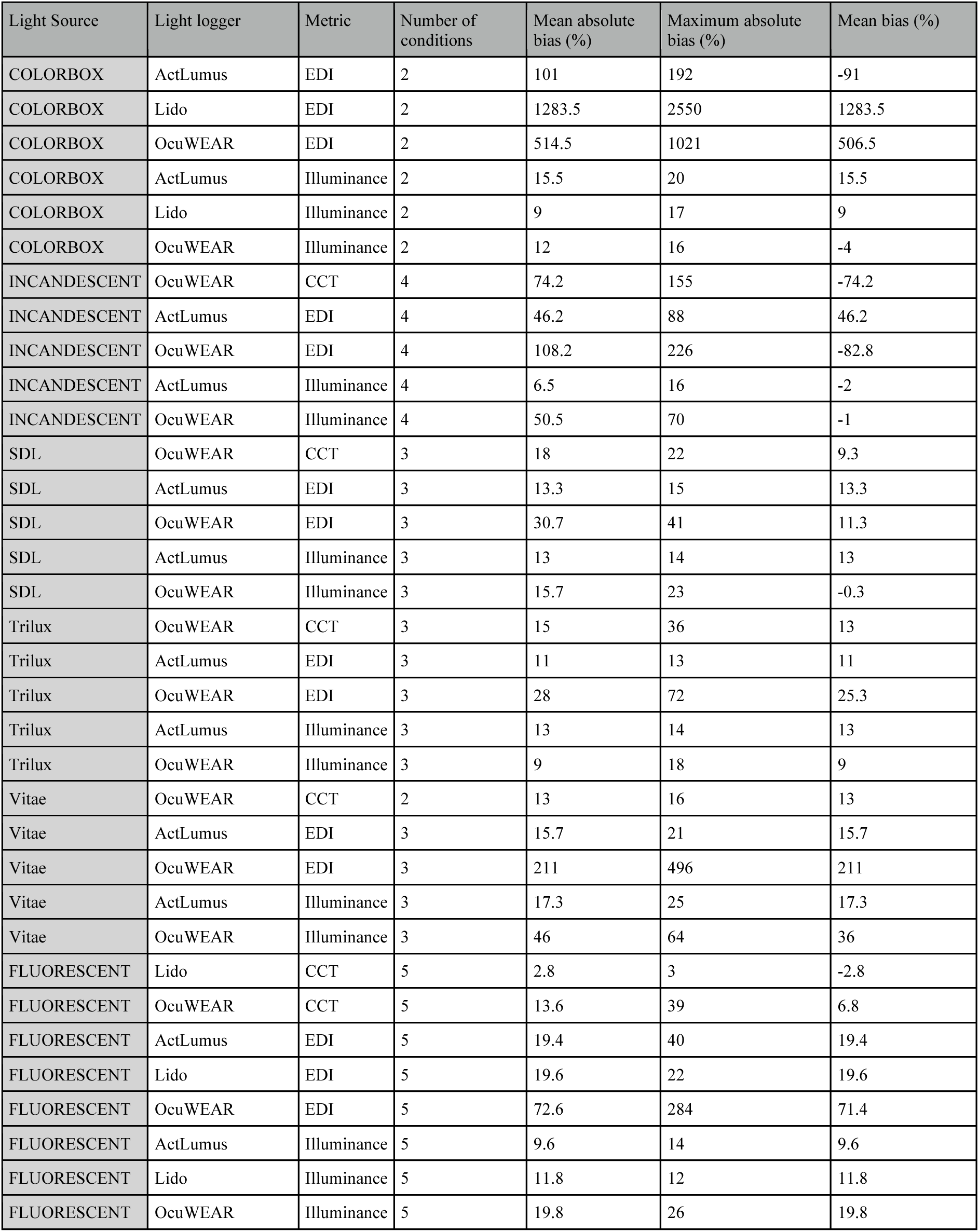
Source-specific performance of wearable light loggers: mean and maximum absolute bias (%) by lighting condition

| Light Source | Light logger | Metric | Number of conditions | Mean absolute bias (%) | Maximum absolute bias (%) | Mean bias (%) |
| --- | --- | --- | --- | --- | --- | --- |
| COLORBOX | ActLumus | EDI | 2 | 101 | 192 | -91 |
| COLORBOX | Lido | EDI | 2 | 1283.5 | 2550 | 1283.5 |
| COLORBOX | OcuWEAR | EDI | 2 | 514.5 | 1021 | 506.5 |
| COLORBOX | ActLumus | Illuminance | 2 | 15.5 | 20 | 15.5 |
| COLORBOX | Lido | Illuminance | 2 | 9 | 17 | 9 |
| COLORBOX | OcuWEAR | Illuminance | 2 | 12 | 16 | -4 |
| INCANDESCENT | OcuWEAR | CCT | 4 | 74.2 | 155 | -74.2 |
| INCANDESCENT | ActLumus | EDI | 4 | 46.2 | 88 | 46.2 |
| INCANDESCENT | OcuWEAR | EDI | 4 | 108.2 | 226 | -82.8 |
| INCANDESCENT | ActLumus | Illuminance | 4 | 6.5 | 16 | -2 |
| INCANDESCENT | OcuWEAR | Illuminance | 4 | 50.5 | 70 | -1 |
| SDL | OcuWEAR | CCT | 3 | 18 | 22 | 9.3 |
| SDL | ActLumus | EDI | 3 | 13.3 | 15 | 13.3 |
| SDL | OcuWEAR | EDI | 3 | 30.7 | 41 | 11.3 |
| SDL | ActLumus | Illuminance | 3 | 13 | 14 | 13 |
| SDL | OcuWEAR | Illuminance | 3 | 15.7 | 23 | -0.3 |
| Trilux | OcuWEAR | CCT | 3 | 15 | 36 | 13 |
| Trilux | ActLumus | EDI | 3 | 11 | 13 | 11 |
| Trilux | OcuWEAR | EDI | 3 | 28 | 72 | 25.3 |
| Trilux | ActLumus | Illuminance | 3 | 13 | 14 | 13 |
| Trilux | OcuWEAR | Illuminance | 3 | 9 | 18 | 9 |
| Vitae | OcuWEAR | CCT | 2 | 13 | 16 | 13 |
| Vitae | ActLumus | EDI | 3 | 15.7 | 21 | 15.7 |
| Vitae | OcuWEAR | EDI | 3 | 211 | 496 | 211 |
| Vitae | ActLumus | Illuminance | 3 | 17.3 | 25 | 17.3 |
| Vitae | OcuWEAR | Illuminance | 3 | 46 | 64 | 36 |
| FLUORESCENT | Lido | CCT | 5 | 2.8 | 3 | -2.8 |
| FLUORESCENT | OcuWEAR | CCT | 5 | 13.6 | 39 | 6.8 |
| FLUORESCENT | ActLumus | EDI | 5 | 19.4 | 40 | 19.4 |
| FLUORESCENT | Lido | EDI | 5 | 19.6 | 22 | 19.6 |
| FLUORESCENT | OcuWEAR | EDI | 5 | 72.6 | 284 | 71.4 |
| FLUORESCENT | ActLumus | Illuminance | 5 | 9.6 | 14 | 9.6 |
| FLUORESCENT | Lido | Illuminance | 5 | 11.8 | 12 | 11.8 |
| FLUORESCENT | OcuWEAR | Illuminance | 5 | 19.8 | 26 | 19.8 |

## References

1. Boivin, D. B., Duffy, J. F., Kronauer, R. E. & Czeisler, C. A. Dose-response relationships for resetting of human circadian clock by light. Nature 379, 540–542 (1996).

2. Khalsa, S. B. S., Jewett, M. E., Cajochen, C. & Czeisler, C. A. A phase response curve to single bright light pulses in human subjects. J. Physiol. 549, 945–952 (2003).

3. Lewy, A. J. Effects of light on human melatonin production and the human circadian system. Prog. Neuropsychopharmacol. Biol. Psychiatry 7, 551–556 (1983).

4. Hilaire, M. A. S. et al. The spectral sensitivity of human circadian phase resetting and melatonin suppression to light changes dynamically with light duration. Proceedings of the National Academy of Sciences 119, e2205301119 (2022).

5. Gubin, D. et al. Blue light and temperature actigraphy measures predicting metabolic health are linked to melatonin receptor polymorphism. Biology (Basel) 13, 22 (2023).

6. Ishihara, A., Courville, A. B. & Chen, K. Y. The Complex Effects of Light on Metabolism in Humans. Nutrients 15, (2023).

7. Cajochen, C., Zeitzer, J. M., Czeisler, C. A. & Dijk, D. J. Dose-response relationship for light intensity and ocular and electroencephalographic correlates of human alertness. Behav. Brain Res. 115, 75–83 (2000).

8. Chellappa, S. L. et al. Non-visual effects of light on melatonin, alertness and cognitive performance: can blue-enriched light keep us alert? PLoS One 6, e16429 (2011).

9. Milosavljevic, N. How does light regulate mood and behavioral state? Clocks Sleep 1, 319–331 (2019).

10. Vandewalle, G., Maquet, P. & Dijk, D.-J. Light as a modulator of cognitive brain function. Trends Cogn. Sci. 13, 429–438 (2009).

11. Dijk, D.-J. & Archer, S. N. Light, sleep, and circadian rhythms: together again. PLoS Biol. 7, e1000145 (2009).

12. Berson, D. M., Dunn, F. A. & Takao, M. Phototransduction by retinal ganglion cells that set the circadian clock. Science 295, 1070–1073 (2002).

13. Hattar, S., Liao, H. W., Takao, M., Berson, D. M. & Yau, K. W. Melanopsin-containing retinal ganglion cells: architecture, projections, and intrinsic photosensitivity. Science 295, 1065–1070 (2002).

14. Provencio, I. et al. A novel human opsin in the inner retina. J. Neurosci. 20, 600–605 (2000).

15. Biller, A. M., Balakrishnan, P. & Spitschan, M. Behavioural determinants of physiologically-relevant light exposure. Commun Psychol 2, 1–11 (2024).

16. Guidolin, C. et al. Protocol for a prospective, multicentre, cross-sectional cohort study to assess personal light exposure. BMC Public Health 24, 1–12 (2024).

17. van Duijnhoven, J. et al. Measuring light exposure in daily life: A review of wearable light loggers. Build. Environ. 112771 (2025).

18. Cajochen, C. Alerting effects of light. Sleep Med. Rev. 11, 453–464 (2007).

19. Agbeshie, G. K. et al. Physiologically relevant real-world light exposure and its behavioural and environmental determinants in Kumasi, Ghana. medRxiv 2025.09.11.25334564 (2025) doi:10.1101/2025.09.11.25334564.

20. Hartmeyer, S. L. et al. Multi-wearable approach for monitoring diurnal light exposure and body rhythms in nightshift workers. Acta Physiol. (Oxf.) 241, e70069 (2025).

21. Stefani, O. et al. Wearable light loggers in field conditions: Corneal light characteristics, user compliance, and acceptance. Clocks Sleep 6, 619–634 (2024).

22. Peeters, S. T., Smolders, K. C. H. J. & de Kort, Y. A. W. What you set is (not) what you get: How a light intervention in the field translates to personal light exposure. Build. Environ. 185, 107288 (2020).

23. Didikoglu, A. et al. Associations between light exposure and sleep timing and sleepiness while awake in a sample of UK adults in everyday life. Proc. Natl. Acad. Sci. U. S. A. 120, e2301608120 (2023).

24. Brown, T. M. et al. Recommendations for daytime, evening, and nighttime indoor light exposure to best support physiology, sleep, and wakefulness in healthy adults. PLoS Biol. 20, e3001571 (2022).

25. Zauner, J., et al. MeLiDos Project Data Hub. (Zenodo, 2026). doi:10.5281/ZENODO.19678543.

26. Webler, F. S., Spitschan, M., Foster, R. G., Andersen, M. & Peirson, S. N. What is the “spectral diet” of humans? Curr Opin Behav Sci 30, 80–86 (2019).

27. Gubin, D. et al. Wearables in Chronomedicine and Interpretation of Circadian Health. Diagnostics 15, 327 (2025).

28. Aarts, M. P. J., van Duijnhoven, J., Aries, M. B. C. & Rosemann, A. L. P. Performance of personally worn dosimeters to study non-image forming effects of light: Assessment methods. Build. Environ. 117, 60–72 (2017).

29. Lucas, R. J. et al. Measuring and using light in the melanopsin age. Trends Neurosci. 37, 1–9 (2014).

30. Ishihara, A. et al. Performance of wearable light sensors for measuring photopic and melanopic illuminance under laboratory and free-living conditions. Sleep 49, zsaf358 (2026).

31. Phillips, A. J. K. et al. High sensitivity and interindividual variability in the response of the human circadian system to evening light. Proc. Natl. Acad. Sci. U. S. A. 116, 12019–12024 (2019).

32. Beygelzimer, A., et al. FNN: Fast nearest neighbor search algorithms and applications. CRAN: Contributed Packages The R Foundation 10.32614/cran.package.fnn (2010).

33. Martin Bland, J. & Altman, D. Statistical methods for assessing agreement between two methods of clinical measurement. Lancet 327, 307–310 (1986).

34. Datta, D. Blandr: Bland-Altman Method Comparison for R, Version 0.6.0. (Zenodo, 2024). doi:10.5281/ZENODO.824514.

35. Bland, J. M. & Altman, D. G. Measuring agreement in method comparison studies. Stat. Methods Med. Res. 8, 135–160 (1999).

36. Cleveland, W. S. & Devlin, S. J. Locally weighted regression: An approach to regression analysis by local fitting. J. Am. Stat. Assoc. 83, 596–610 (1988).

37. Carkeet, A. Exact parametric confidence intervals for Bland-Altman limits of agreement. Optom. Vis. Sci. 92, e71–80 (2015).

38. Rea, M. S., Figueiro, M. G., Bullough, J. D. & Bierman, A. A model of phototransduction by the human circadian system. Brain Res. Brain Res. Rev. 50, 213–228 (2005).

39. Prins, N. & Frederick, A. A. Psychophysics. (Academic Press, San Diego, CA, 2016).

40. Stevens, S. S. On the psychophysical law. Psychol. Rev. 64, 153–181 (1957).

